# Strategies that make vaccination easy and promote autonomy could increase COVID-19 vaccination in those who remain hesitant

**DOI:** 10.1101/2021.05.19.21257355

**Authors:** Ingrid Eshun-Wilson, Aaloke Mody, Khai Hoan Tram, Cory Bradley, Alexander Sheve, Branson Fox, Vetta Thompson, Elvin H Geng

**Author notes:** Corresponding author: Ingrid Eshun-Wilson, 4523 Clayton Ave, CB 8051, St. Louis MO, 63110, USA, 7073386621. Competing Interest Statement: Authors do not have any competing interests.

## Abstract

The COVID-19 vaccination campaign in the US has been immensely successful in vaccinating those who are receptive, further increases in vaccination rates however will require more innovative approaches to reach those who remain hesitant, deliberative or indifferent. Phenomena such empty mass vaccination sites and wasted vaccine doses in some regions suggest that in addition to dispelling misinformation and building trust, developing more person-centered vaccination strategies, that are modelled on what people want could further increase uptake. To inform vaccine distribution strategies that are aligned with public preferences for COVID-19 vaccination campaign features we conducted a survey and discrete choice experiment among a representative sample of 2,895 people in the US, between March 15 and March 22, 2021. We found that on average the public prioritized ease, preferring single to two dose vaccinations, vaccinating once rather than annually and reduced waiting times at vaccination sites - for some these were the primary preference drivers. Vaccine enforcement reduced overall vaccine acceptance, with a trend of increasing ‘control aversion’ with increasing vaccine hesitancy, particularly among those who were young, Black/African American or Republican. These data suggest that making vaccination easy and promoting autonomy by offering the public choices of vaccination brands and locations may increase uptake, and that vaccine mandates could compromise autonomy and increase control aversion in those who are hesitant - reducing vaccination in such groups and potentially undermining the goals of COVID-19 vaccination campaigns.

**Significance:** DCE’s are a novel tool in public health that allow examination of preferences for a product, service or policy, identifying how the public prioritizes personal risks and cost in relation to health behaviors. Using this method to establish preferences for COVID-19 vaccination campaign strategies, our results suggest that: firstly, vaccination should be made as easy as possible for the public, second, that individuals should be offered choices of vaccine brand and vaccination location, and third, that vaccine enforcement could activate ‘control aversion’ in the public and particularly in those who are most hesitant – potentially causing these groups to double down on vaccine resistance, a scenario which would impede the success of vaccination programs in the US.

## Introduction

The roll out of the COVID-19 vaccine in the United States faces represents a new phase in the COVID-19 response, but must confront an old challenge to public health efforts in this country: the success of biomedical research and the availability of efficacious interventions does not mean that all segments of the public, even as access expands, are interested and motivated to use the intervention. COVID-19 vaccine, in particular, faces numerous barriers to reach. Some in the public are skeptical of the safety of a vaccine developed rapidly. Others are unconvinced that COVID-19 represents a serious health problem. Finally, COVID-19 has been uniquely politicized, and membership in a social group or political persuasion intersect with vaccine reluctance and rejection. Since the problem is not merely one of individual health outcomes, but of societal health and function in general, some scholars have framed the problem of vaccine uptake as a tension between enforcement, incentives vs. prosocial volunteerism. Under these conditions of widespread uncertainty or doubt, the features of a national vaccine program must be aligned with preferences both for the mundane (e.g., wait times) as well as meaningfulness (e.g., the signification of enforcement) to the extent possible to achieve optimal uptake.

While the public health research has used surveys to capture the attitudes (either for or against vaccination), knowledge of how different public health strategies and program attributes will influence preferences, tradeoffs between different desired attributes, as well as how these preferences are related to socio demographic groups is poorly understood. Some experts have advocated for vaccine passports that allow access to desired places (e.g., travel) (1), but others have warned that mandates and restrictions can lead to backlash. In addition, given that preferences are often segmented (different groups have different desires) and that these preference groups do not fall along obvious sociodemographic lines (e.g., age, sex, race), research to reveal different preference phenotypes is also lacking. Finally, while research has also focused on vaccine characteristics that are most desirable (i.e. durability and efficacy), these studies have not drawn attention to the elements of delivery services. In short, information about where, how, when and with what inducements vaccines should best be offered remains less than crystal clear.

Rational utility theory suggests that humans are utility (sometimes translated as happiness) maximizers, and that given constraints, we consider the attributes of goods and services that maximize the utility derived. While embellished by behavioral economics, this notion remains one pillar of behavioral sciences. Discrete choice experiments are a method for soliciting utilities associated with different versions of a good or service. In short, by offering a respondent a choice between repeated versions of a good or service composed of different characteristics within a set attributes (product or service features), a choice experiment can reveal what elements of a good or services are preferred, how strong that preference is (in relation to another), as well the size and characteristics of sub-groups in a population with shared preferences. Discrete choice experiments have been a mainstay of marketing research for decades, and have recently become more widely used in public health to discern the voice of the “consumer.” We report on a nationally representative discrete choice experiment on attributes of vaccination campaigns for the COVID-19 pandemic to inform the design of vaccine program and policy.

## Materials and Methods

We followed the ISPOR guidelines for design of choice experiments (2, 3).

### Choice experiment attribute and attribute level selection

We explored the literature and followed attribute selection guidelines to first generate a comprehensive list of attributes based on expert interviews, discussion and literature, then conduct data reduction, removal of inappropriate attributes and wording refinement (4, 5). Because of evidence of vaccine hesitancy in minority groups and to ensure community participation in the design of the choice experiments we reached out to community leaders as identified by the Center for Community Health Partnership and Research at the Institute for Public Health at Washington University to provide input on the attribute list and levels. We then developed a final set of salient, plausible, manipulable, non-dominant attributes that represented as complete as possible a list of vaccination campaign features (Table 3). We specifically excluded vaccine safety and efficacy due to the dominance of these attributes over implementation concerns seen in other COVID-19 choice experiments.

### Choice experiment design

We limited the number of attributes in the DCE according to design guidelines (seven attributes) and selected those attributes which we determined to be key decision drivers and of the greatest public health policy significance during the time period. To balance statistical and response efficiency (avoid fatigue in respondents) we opted for 2 scenarios and limited the number of attribute levels and prohibited attribute level combinations, we further limited the number of DCE questions asked of each respondent to 10, we opted for two scenarios per task. We manually removed combinations considered non sensical (drop-in scheduling combined with waiting time of 1 or 2 hours). Participants chose between two vaccination scenarios and to prevent forced responses we included a third opt-out (choose neither) scenario. The attribute levels were randomly ordered across participants and each participant received one of 300 versions of the choice experiment. The survey was designed in Lighthouse studio, Sawtooth software, a choice experiment design tool (6). We generated a near-orthogonal (each pair of attribute levels appears equally across the experiment) and near-balanced (each level appears equally often across the experiment), fractional factorial design. We assumed there were no interactions between attributes. The choice tasks were randomly ordered between participants to prevent bias induced by question order. To later assess internal validity and comprehension of the experiment we included a within-set dominant pair: a fixed task question where 6 attribute levels were kept constant and as desirable as possible (health facility vaccination location, almost all in your community vaccinated, no vaccination enforcement, 1 dose and only 1 vaccination needed) and 1 attribute which clearly dominated the other - 2 hours waiting time versus no waiting time (7). Sample size estimation: Although routine choice experiment sample sizes can be estimated using the formula N ≥ (500 × c)/(a × t) - where N is the number of participants, t is the number of choice tasks (questions), a is the number of alternative scenarios and c is the largest number of attribute levels for any one attribute (8), the inclusion of prohibited attribute level combinations in our design resulted in our use of the Sawtooth logit efficiency test which allows for the evaluation of sample sizes against simulated utility estimates to ensure utility standard errors remained <0.05.

### Data collection and measurements

In order to conduct subgroup analyses we additionally collected data on demographic characteristics including: age, gender, race group, employment status, political affiliation. We also collected data on COVID-19 vaccination status, and intention to vaccinate based on questions from the Census pulse survey (9). The survey tool and DCE tool can be found in Appendix S1. We recruited participants through Qualtrics Research Services (10). Qualtrics is a recognized leader in survey recruitment methodology, utilizing multiple consumer panels to obtain nationally representative survey samples. We used this approach to ensure sufficient samples subgroups by age, gender, race and Hispanic/Latino ethnicity, to ensure both a nationally representative sample and by oversampling particular groups to allow for later choice experiment subgroup analyses by ensuring sufficient participants (more than 200) in each age group, gender group, race (Black/African American and white) and Hispanic/Latino subgroups. Participants were recruited online and offered incentives based on their profile within the Qualtrics system which varied by participant (e.g. cash, gift cards, airmiles). Prospective survey participants were sent email invitations or prompted on the survey platform to proceed with a given survey. The survey was initially piloted in 10 participants who provided feedback on the design and subsequently a further 100 anonymous participants, after reviewing the quality of responses and feedback the survey was further refined. To improve comprehension of the DCE an example of a simplistic choice experiment was presented to participants prior to eliciting responses to the 10 choice tasks.

### Analyses

We first generated population inverse probability sampling weights to reweight our sample to reflect the US Census population estimates for age, race, gender and vaccination status or intention – (Census Pulse Survey 27: March17 - March 29, 2021) from a time frame which corresponded to the period our survey was conducted (9). Census population proportions as applied to weights are presented in Table S5.

Choice experiment modelling is based on random utility theory (RUT) which assumes that the utility (U) for individual i conditional on choice j consists of an explainable component (Vij) and a random component (e_ij_) (formula 1). The random component may capture any combination of unobserved attributes, unobserved preference variation, specification error, measurement error and inherent variability within and between individuals (11).

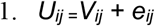

For this analysis we applied dummy coding and created an alternative specific constant to represent the opt-out choice scenario (12, 13). For main effects we conducted mixed logit regression models to account for preference heterogeneity with all attributes included as random parameters and the opt-out ASC included as a fixed parameter. The explainable component (V_ij_) for this experiment is denoted in formula 2 below, where b_1-15_ represents the coefficient for the corresponding attribute level and b_o_ represents the utility associated with the ASC. The baseline attribute category for each attribute is omitted from formulae and estimations, as this attribute has by definition a utility of 0 when dummy coding is used.

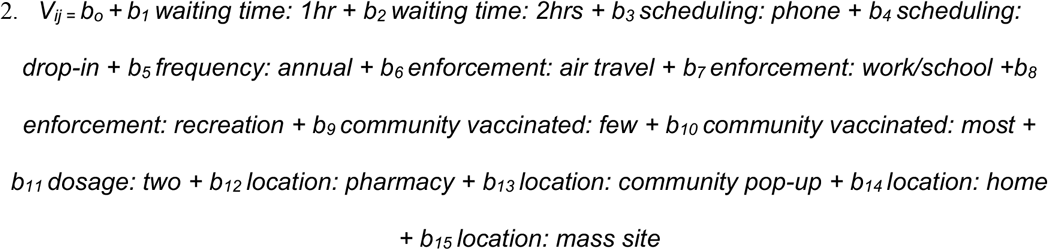

In this analysis the utility for the ASC includes the utility for the opt-out choice, inattention, experiment complexity and is confounded by the utility estimates for the baseline levels of attributes, b_o_ can therefore not be directly interpreted from model outputs. For this choice experiment our interest was relative utilities and no estimation of preference shares or probability of uptake, we therefore used dummy coded datasets in estimations (14). Mixed logit models were fit using Stata’s mixlogit command which uses simulated maximum likelihood estimators and generates mean utilities for the population and standard deviations of the random coefficients (15). Mixed logit coefficients (b) can be interpreted as the strength of the relative preference for the particular attribute comparison, with positive coefficients representing positive preferences (desirable) and negative coefficients representing negative preferences (less desirable). Standard deviations represent preference heterogeneity for attribute comparisons, with a 0 standard deviation indicating no heterogeneity.

We fit up to seven latent class conditional logit models using maximum likelihood estimation of datasets expanded by sampling weights and selected the model with the smallest model fit criterion (Akaike and Bayesian information criterion), the highest mean probability of group membership and the smallest number of participants with a low probability of group membership in each group. We additionally qualitatively evaluated latent classes. We validated latent class membership using cross-validation techniques (16). We used multinomial logistic regression models and relative risk ratios with 95% confidence intervals to evaluate predictors of latent class membership firstly for participant demographic characteristics and secondly for vaccination status or intention and additionally present marginal probabilities of latent class membership based on these models.

The study was approved by the Washington University institutional review board and was exempted from a formal consent process, participants were provided with an introductory information sheet as part of the online survey.

## Results

### Participant characteristics

The survey was fielded between March 15, 2021 and March 22, 2021 (Figure 1) and participants characteristics are presented in Table 1. Of 2,985 survey respondents 38%were already vaccinated (received at least one dose), (41.6%) would definitely get vaccinated, 28.6% would probably get vaccinated, 18.4% would probably not get vaccinated and 9.8% would definitely not get vaccinated. Participants were located in all 50 US states, as well as the District of Columbia, 89.0% resided in an urban setting and 11.0% in rural areas. Men made up 37.2% of the sample, with the remaining reporting female gender. The median age was 50 years with an interquartile range (IQR) of 33 to 62 years. 23.5% of the sample was Black/African American, 64.1% White, 7.9% Asian and 4.5% other race groups, and 14.8% identified as Hispanic/Latina.

**Figure 1:**
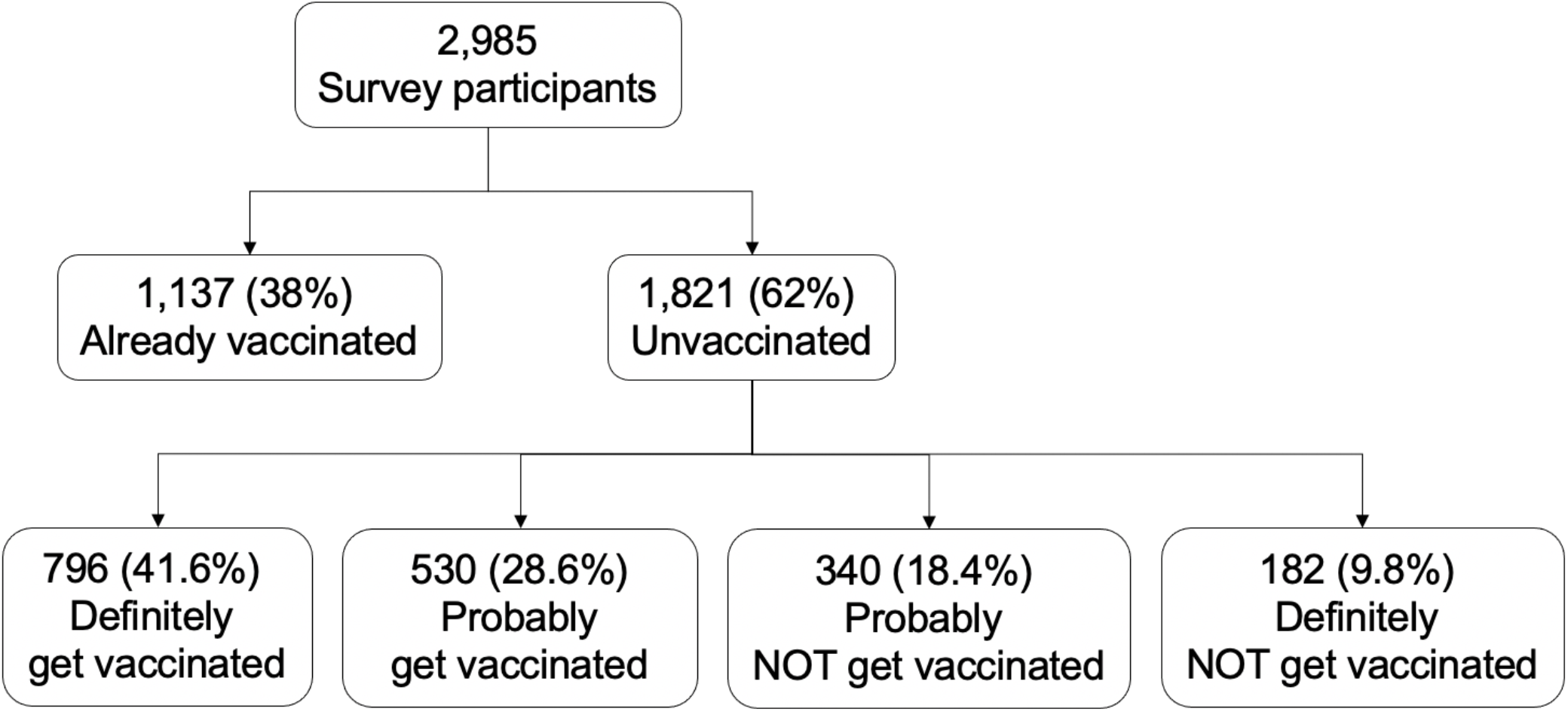
Survey flow diagram for analysis sample. Numbers are unweighted. Survey was delivered between March 15 to March 22, 2021.

**Figure 2:**
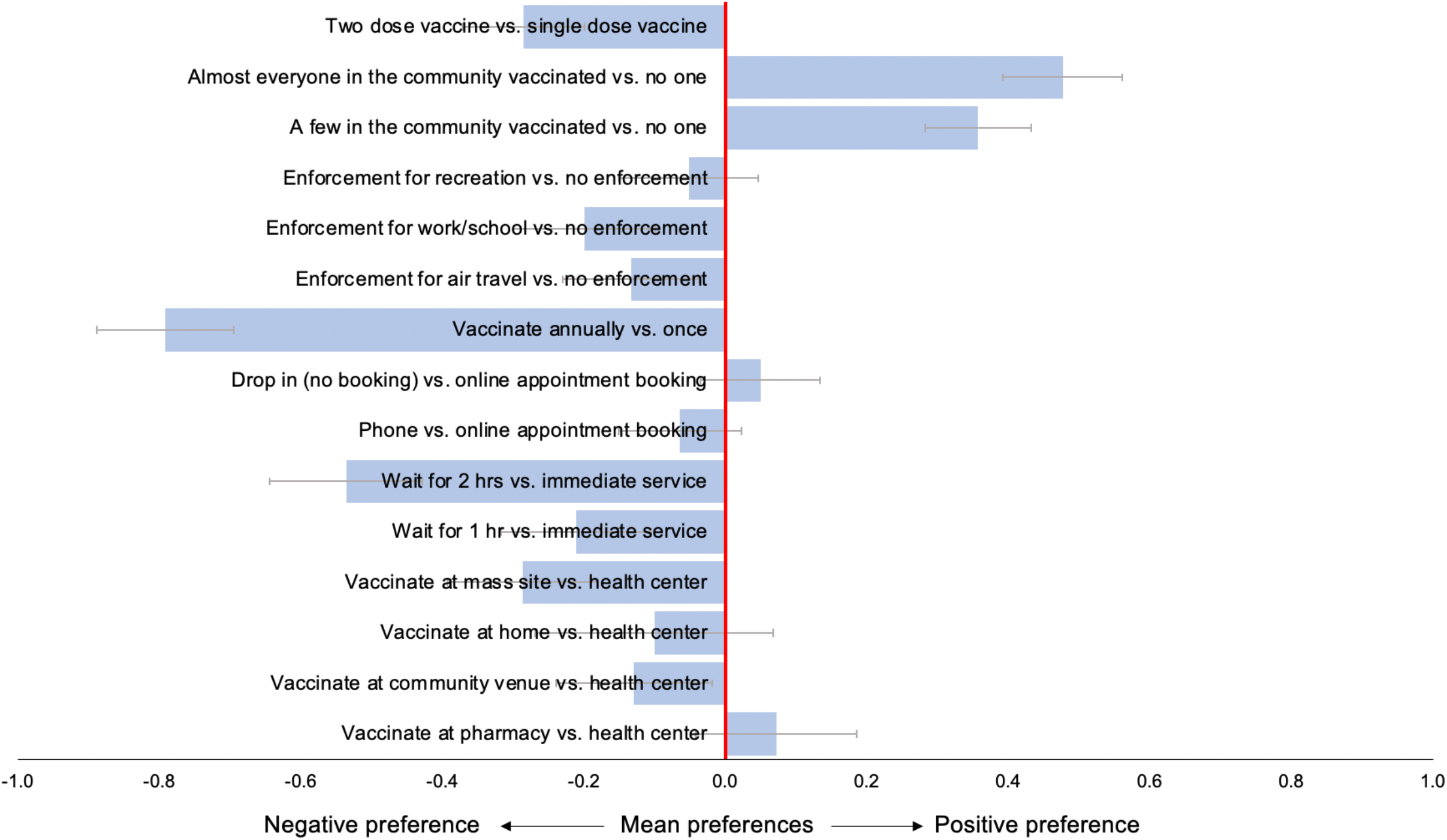
Weighted mean preferences (relative utilities) for vaccination campaign features, in total population (N=2,985)

**Table 1:** Characteristics and COVID experiences of participants.

| Participant characteristics |  | Total<br>(N=2,985) | Already<br>vaccinated<br>(N=1,137) | Definitely<br>get a<br>vaccine<br>(N=796) | Probably get<br>a vaccine<br>(N=530) | Probably<br>NOT get a<br>vaccine<br>(N=340) | Definitely<br>NOT get a<br>vaccine<br>(N=182) | p-value |
| --- | --- | --- | --- | --- | --- | --- | --- | --- |
| Age (years) – median; IQR |  | 50.0 (33.0-62.0) | 60.0 (41.0-68.0) | 46.0 (34.0-58.0) | 41.0 (26.0-55.0) | 43.0 (27.0-56.0) | 37.5 (22.0-55.0) | <0.001 |
| Male gender |  | 1,111 (37.2%) | 448 (39.4%) | 329 (41.3%) | 175 (33.0%) | 101 (29.7%) | 58 (31.9%) | <0.001 |
| Hispanic/Latino ethnicity |  | 443 (14.8%) | 142 (12.5%) | 122 (15.3%) | 95 (17.9%) | 52 (15.3%) | 32 (17.6%) | 0.036 |
| Race | Black/African American | 225 (19.8%) | 702 (23.5%) | 155 (19.5%) | 156 (29.4%) | 107 (31.5%) | 59 (32.4%) | <0.001 |
|  | White | 808 (71.1%) | 1,913 (64.1%) | 508 (63.8%) | 299 (56.4%) | 191 (56.2%) | 107 (58.8%) |  |
|  | Asian | 65 (5.7%) | 235 (7.9%) | 99 (12.4%) | 47 (8.9%) | 18 (5.3%) | 6 (3.3%) |  |
|  | Other* | 135 (4.5%) | 39 (3.4%) | 34 (4.3%) | 28 (5.3%) | 24 (7.1%) | 10 (5.5%) |  |
| Schooling | High school or less | 189 (16.6%) | 712 (23.9%) | 180 (22.6%) | 158 (29.8%) | 109 (32.1%) | 76 (41.8%) | <0.001 |
|  | Incomplete college/Associate degree | 354 (31.1%) | 993 (33.3%) | 252 (31.7%) | 187 (35.3%) | 136 (40.0%) | 64 (35.2%) |  |
|  | Bachelor degree or higher | 594 (52.2%) | 1,280 (42.9%) | 364 (45.7%) | 185 (34.9%) | 95 (27.9%) | 42 (23.1%) |  |
| Political affiliation | Democrat | 605 (53.2%) | 1,425 (47.7%) | 439 (55.2%) | 216 (40.8%) | 103 (30.3%) | 62 (34.1%) | <0.001 |
|  | Republican | 291 (25.6%) | 748 (25.1%) | 155 (19.5%) | 137 (25.8%) | 111 (32.6%) | 54 (29.7%) |  |
|  | Other | 67 (5.9%) | 222 (7.4%) | 48 (6.0%) | 58 (10.9%) | 35 (10.3%) | 14 (7.7%) |  |
|  | Unaffiliated | 156 (13.7%) | 476 (15.9%) | 128 (16.1%) | 85 (16.0%) | 70 (20.6%) | 37 (20.3%) |  |
|  | Prefer not to answer | 18 (1.6%) | 114 (3.8%) | 26 (3.3%) | 34 (6.4%) | 21 (6.2%) | 15 (8.2%) |  |
| Setting | Urban | 1,020 (89.7%) | 2,656 (89.0%) | 700 (88.1%) | 482 (90.9%) | 290 (85.3%) | 164 (90.1%) | 0.079 |
|  | Rural | 117 (10.3%) | 328 (11.0%) | 95 (11.9%) | 48 (9.1%) | 50 (14.7%) | 18 (9.9%) |  |
| Any children |  | 1,664 (55.7%) | 737 (64.8%) | 408 (51.3%) | 274 (51.7%) | 164 (48.2%) | 81 (44.5%) | <0.001 |
| Worked for pay/profit in the last 7 days |  | 1,457 (48.8%) | 566 (49.8%) | 388 (48.7%) | 256 (48.3%) | 168 (49.4%) | 79 (43.4%) | 0.620 |
| Reason for not working in the last 7 days | Retired or do not want to be employed | 732 (47.9%) | 412 (72.2%) | 151 (37.0%) | 80 (29.2%) | 54 (31.4%) | 35 (34.0%) | <0.001 |
|  | Sick or caring for others -COVID related | 109 (7.1%) | 17 (3.0%) | 33 (8.1%) | 31 (11.3%) | 19 (11.0%) | 9 (8.7%) |  |
|  | Sick or caring for others - not COVID related | 170 (11.1%) | 46 (8.1%) | 44 (10.8%) | 44 (16.1%) | 24 (14.0%) | 12 (11.7%) |  |
|  | I was concerned about getting or spreading the coronavirus | 74 (4.8%) | 15 (2.6%) | 35 (8.6%) | 15 (5.5%) | 6 (3.5%) | 3 (2.9%) |  |
|  | Job loss due to COVID economic impact | 176 (11.5%) | 35 (6.1%) | 63 (15.4%) | 33 (12.0%) | 32 (18.6%) | 13 (12.6%) |  |
|  | Other reason | 267 (17.5%) | 46 (8.1%) | 82 (20.1%) | 71 (25.9%) | 37 (21.5%) | 31 (30.1%) |  |
| **Applied for UI benefits** |  | 628 (21.0%) | 264 (23.2%) | 137 (17.2%) | 122 (23.0%) | 70 (20.6%) | 35 (19.2%) | 0.018 |
| **Anyone in the family lost income since Mar13, 2020** |  | 1,117 (37.4%) | 391 (34.4%) | 309 (38.8%) | 220 (41.5%) | 133 (39.1%) | 64 (35.2%) | 0.045 |
| **Anyone in the family likely to lose income due to COVID-19 next 4wks** |  | 581 (19.5%) | 224 (19.7%) | 150 (18.8%) | 108 (20.4%) | 68 (20.0%) | 31 (17.0%) | 0.87 |
| **Previous COVID-19 infection** | Yes | 198 (17.4%) | 397 (13.3%) | 78 (9.8%) | 73 (13.8%) | 33 (9.7%) | 15 (8.2%) | <0.001 |
|  | No | 932 (82.0%) | 2,534 (84.9%) | 711 (89.3%) | 442 (83.4%) | 292 (85.9%) | 157 (86.3%) |  |
|  | Unsure | 7 (0.6%) | 54 (1.8%) | 7 (0.9%) | 15 (2.8%) | 15 (4.4%) | 10 (5.5%) |  |
| **Eligible to receive a COVID-19 vaccine?** | Yes | 1,716 (57.5%) | 1,137 (100.0%) | 261 (32.8%) | 158 (29.8%) | 100 (29.4%) | 60 (33.0%) | <0.001 |
|  | No | 928 (31.1%) | 0 (0.0%) | 456 (57.3%) | 258 (48.7%) | 137 (40.3%) | 77 (42.3%) |  |
|  | Unsure | 341 (11.4%) | 0 (0.0%) | 79 (9.9%) | 114 (21.5%) | 103 (30.3%) | 45 (24.7%) |  |
| **Reason for not vaccinating yet, among eligible who intend to vaccinate** | I tried but could not get an appointment |  |  | 118 (45.2%) | 46 (29.1%) |  |  | 0.004 |
|  | The vaccination site is too far away |  |  | 18 (6.9%) | 21 (13.3%) |  |  |  |
|  | I could not find information on where to get a vaccination |  |  | 25 (9.6%) | 23 (14.6%) |  |  |  |
|  | Other reason |  |  | 100 (38.3%) | 68 (43.0%) |  |  |  |
| **Reasons for hesitancy among those probably/definitely not going to vaccinate\*\*** | Deliberative reasons | 407 (78%) |  |  |  | 278 (81.8%) | 129 (70.9%) | 0.004 |
|  | Dissenting reasons | 103 (19.7%) |  |  |  | 63 (18.5%) | 40 (22.0%) | 0.350 |
|  | Distrustful reasons | 198 (37.9%) |  |  |  | 112 (32.9%) | 86 (47.3%) | 0.001 |

\*Other race includes: American Indian (N=50); Native Hawaiian (N=8); Filipino (N=39); Samoan (N=1) and other Pacific Islander (N=37)
\*\**Deliberative reasons*: I am concerned about the cost of the vaccine; I think other people need it more; I plan to wait and see if it is safe; My doctor has not recommended it; I don't know if the vaccine will work; I am concerned about possible side effects; I already had COVID-19; I am not a member of a high-risk group; I plan to use masks or other precautions instead. *Distrustful reasons*: I don't trust the government; I don't trust COVID-19 vaccines. *Dissenting reasons*: I don't like vaccines; I don't believe COVID-19 is a serious illness; I don't think vaccines are beneficial.

Those who were eligible to vaccinate and intended to vaccinate but had not received vaccinations at the time of the study, reported struggles to secure vaccine appointments (39%), distance from vaccination sites (9.3%) and lack of information on vaccination locations (11.5%) as reasons for remaining unvaccinated. Those who probably or definitely did NOT intend to vaccinate most commonly reported deliberative reasons for not vaccinating such as concerns about side-effects and efficacy (78.0%), followed by distrustful reasons including mistrust of the government or COVID vaccines (37.9%), and lastly dissenting reasons including an overarching lack of belief in vaccines or that COVID-19 is a serious illness (19.7%).

### Main preferences

In the entire population (N=2,985), weighted preference estimates for COVID-19 vaccination campaign features (Figure 1, Table S1) demonstrated that compared to vaccination at a health facility, the US public were equally willing to receive vaccinations at pharmacies (mean preference 0.07; 95%CI: -0.04 to 0.19) or at home (mean preference -0.10; 95%CI: -0.27 to 0.07), with a slight negative preference for community venues (relative utility: -0.13; 95%CI: -0.24 to -0.02) and a stronger negative preference for mass vaccination sites supported by national guard (mean preference -0.29; 95%CI: -0.39 to -0.18). Immediate service was preferred to longer waiting times of 1 hour (mean preference -0.21; 95%CI: -0.32 to -0.11) or 2 hours (mean preferences -0.54; 95%CI: -0.64 to -0.43). Phone appointment scheduling (mean preference -0.07; 95%CI: -0.19 to 0.05) and drop-in appointments (mean preference 0.05; 95%CI: -0.03 to 0.13) were equally preferred to online appointment scheduling. When compared to no enforcement, preferences for enforcement measures were consistently negative, for air travel (mean preference -0.13; 95%CI: -0.23 to -0.04), work or school attendance (mean preference -0.20; 95%CI: -0.30 to -0.10) or group recreational activities (mean preference -0.05; 95%CI: -0.15 to 0.05), with vaccination enforcement for group recreational activities being the most acceptable compared to work or air travel enforcement. Participants preferred vaccination scenarios where a few people (mean preference 0.36; 95%CI: 0.28 to 0.43) or almost everyone (mean preference 0.48; 95%CI: 0.39 to 0.56) in their community were already vaccinated compared none. Two vaccination doses were less preferred than a single vaccination dose (mean preference -0.29 to -0.37 to -0.20). The strongest preference across attributes was a negative preference for annual vaccinations compared to one vaccination episode for long-term immunity (mean preference 0.79; 95%CI: -0.89 to -0.70). There was substantial preference heterogeneity, as evidenced by large standard deviations (SD) for several attributes generated by the mixed logit model (Table S1), in particular for vaccination at home (SD 0.93, 95%CI: 0.74-1.13), annual vaccination (SD 1.07; 95%CI: 0.97 to 1.18) and number of vaccination doses required (SD 0.97, 95%CI: 0.83. to 1.10).

### Preferences by vaccination status and intention

Subgroup analyses by vaccination status and intention showed largely similar preferences among vaccinators, the already vaccinated or would “definitely get vaccinated” (Figure 3(a), Table S2), mirroring the main population preferences, with the exception that those who had already vaccinated had a slightly stronger preference for vaccinating only once for long-term immunity (mean preference “already vaccinated” – 0.96; 95%CI: -1.13 to -0.80, “definitely get vaccinated” -0.69; 95%CI: -0.85 to -0.53) and were more accepting of a two-dose vaccine (mean preference “already vaccinated” – 0.13; 95%CI: -0.30 to 0.03, “definitely get vaccinated” -0.41; 95%CI: -0.55 to -0.27).

**Figure 3:**
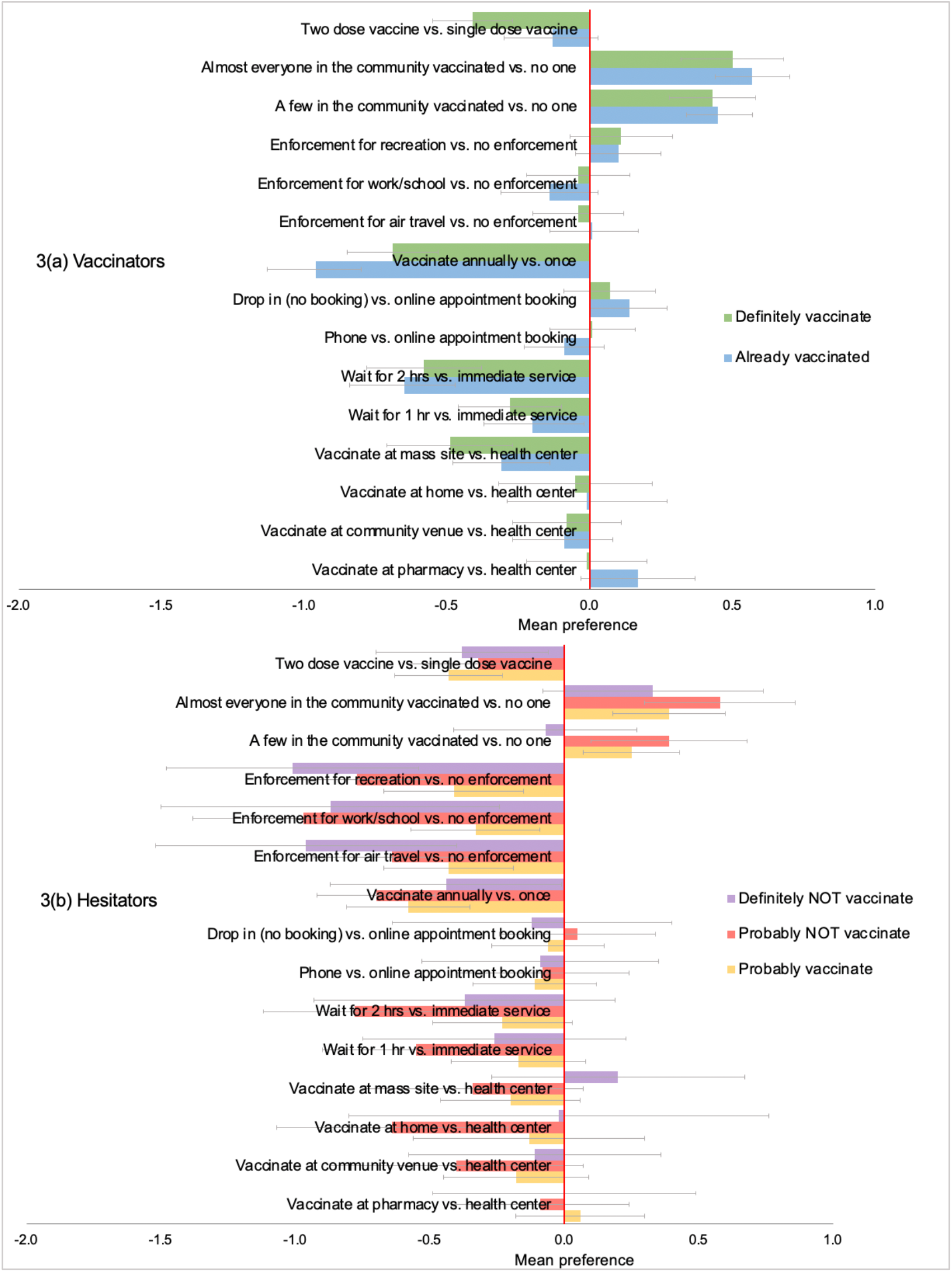
Weighted mean preferences (relative utilities) for vaccination campaign features, among (a) vaccinators and (b) hesitators.

Mean preferences among hesitators, those who would “probably get vaccinated”, “probably NOT get vaccinated” or “definitely NOT get vaccinated” (Figure 3(a), Table S2) appeared similar to main preferences for waiting time, appointment scheduling, vaccination frequency and number of vaccine doses. For vaccination enforcement however, those who were more hesitant were less willingness to vaccinate under enforcement, with increasingly negative preferences towards enforcement with increasing levels of hesitancy (mean preferences for enforcement for work or school: “probably get vaccinated” -0.33; 95%CI -0.56 to -0.09, “definitely NOT get vaccinated” -0.87; 95%CI: -1.50 to -0.24). In addition, those who stated they would “definitely NOT vaccinate” were less influenced by vaccination coverage in the community than other hesitant groups, indicating that vaccination coverage in their community would not influence their decision to vaccinate if only a few were vaccinated (mean preference: -0.07; 95%CI: -0.41 to 0.27), and would only slightly influence them if almost everyone was vaccinated (mean preference: 0.33; 95%CI: -0.08 to 0.74). Those with less hesitancy, who would ‘probably get vaccinated’ or ‘probably NOT get vaccinated’ preferred to vaccinate when at least a few in the community were vaccinated with little difference between whether a few (mean preference among probably NOT get vaccinated: 0.39; 95%CI: 0.10 to 0.67) or almost all of their community was vaccinated (mean preference: 0.20 among probably NOT get vaccinated: 0.58; 95%CI: 0.29 to 0.86). There was some heterogeneity of preferences for vaccination location when compared to health facility vaccination among vaccine hesitant groups, with those who would “probably NOT get vaccinated” showing a negative preference for home vaccination (mean preference: -0.64; 95%CI: -1.08 to -0.21) and those who would definitely NOT get vaccinated finding mass vaccination sites more acceptable than other groups (mean preference: 0.20; 95%CI -0.27 to 0.67).

### Latent class analysis

Latent class analysis revealed six distinct preference subgroups in the population (Figure 4(a-d), Table S3). Approximately half of the population (46.1%) belonged to one of three latent class preference groups who were more concerned about inherent vaccine features (Fig 4a) such as dosage and frequency than vaccine distribution approaches: this included a “single dose” group (7.9%) who showed a strong negative preference for two vaccine doses (mean preference: -4.21; 95%CI: -4.90 to -3.53), a “two dose” group (15.8%) whose strongest preference was for two vaccine doses instead of one (mean preference: 1.72; 95%CI: 1.49 to 1.95) and a “vaccinate once” group (22.4%) whose strongest preference was for a single vaccination episode offering long-term immunity rather than annual vaccination (mean preference: -3.88; 95%CI: -4.18 to -3.57).

**Figure 4 (a-f):**
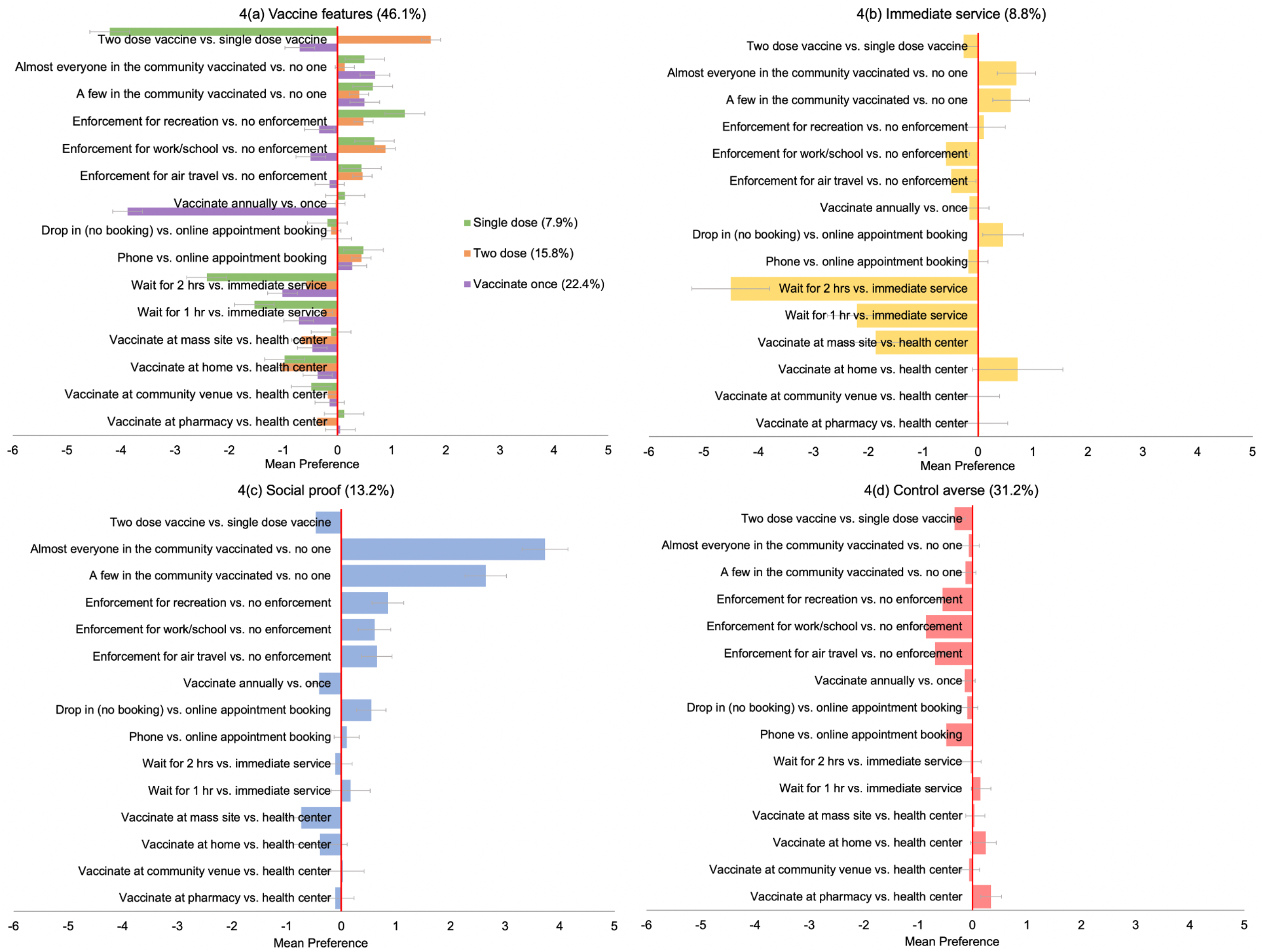
Weighted mean preferences (relative utilities) for vaccination campaign features, by latent class membership group.

The fourth group were focused on services at the vaccination site, with strong negative preferences for waiting time of one hour (−2.21; 95%CI: -2.75 to -1.67) or two hours (mean preference: -4.51; 95%CI: -5.21 to - 3.80) and the strongest negative preference across groups for mass vaccination sites compared to vaccination at the health facility (mean preference: -1.87; 95%CI: -2.55 to -1.19) (Figure 4d).

The sixth preference subgroup, the “social proof” group (13.2%) (Fig 4e) were the only group to show primarily positive preferences indicating what would increase their probability of vaccination, this group had a strong preference for a few in their community to be vaccinated first rather than no-one (mean preference 2.64; 95%CI: 2.26 to 3.02), their preference for vaccination increased with greater vaccine coverage (mean preference 3.73; 95%CI: 3.31 to 4.15). They additionally showed relatively weak positive preferences for vaccination enforcement (mean preference air travel: 0.65; 95%CI: 0.37 -0.93).

The final and largest preference subgroup, the “control averse” group (31.9%) (Fig 4f) were generally indifferent to inherent vaccine features and distribution strategies but showed consistently negative preferences for vaccination enforcement, for work or school (mean preference: -0.86; 95% CI: -1.03 to -0.69), air travel (mean preference: -0.69; 95%CI: -0.86 to -0.52) or recreation (mean preference: -0.55; 95%CI: -0.71 to -0.3

### Predictors of latent class membership

Those who were affiliated with the republican party were more likely to be driven by preferences for immediate service (RRR 1.73; 95%CI: 1 to 2.99), specific vaccine features (RRR 2.36; 95%CI: 1.51 to 3.69), or control aversion (RRR 2.16; 95%CI: 1.38 to 3.37), compared to those who desired ‘social proof’ of vaccine safety (Table 2). Black/African Americans were twice as likely as whites to be control averse rather than in the ‘social proof’ group (RRR 2.00; 95%CI: 1.28 to 3.12) and were more concerned about inherent vaccine features (RRR 1.63: 95%CI: 1.05 to 2.53). Older age was also a driver of belonging to latent class groups that prioritized immediate service (RRR 1.16; 95%CI: 1.28 to 3.12) or vaccine features (RRR 2.00; 95%CI: 1.28 to 3.12), when compared to those in ‘social proof’ subgroup. The marginal probability of being in the ‘social proof’ group (Figure 5) was 15.2% for democrats and 7.5% for republicans, 7.9% for black/African Americans, 13.1% for whites and 15.5% for Asians, 17.6% for those 18 to 24 years old and 9.6% for those who were over 65years. The marginal probability of being ‘control averse’ was 33.7% for republicans and 31.8% for democrats, 38.4% for Black/African Americans, 31.5% for whites, 25.5% for Asians and 43.5% for other race groups; 42.7% for those 25-34 years and 20.1% for those over 65 years (Figure 5, Table S4). Table 2: Weighted multinomial logit model: Predictors of latent class membership for the “control averse”, “vaccine feature” and “immediate service” preference groups relative to the “social proof” preference group (13.2%).

**Table 2:** Weighted multinomial logit model: Predictors of latent class membership for the “control averse”, “vaccine feature” and “immediate service” preference groups relative to the “social proof” preference group (13.2%).

| Participant characteristics |  | Immediate service (8.8%) |  |  |  | Vaccine features (47.9%) |  |  |  | Control averse (31.9%) |  |  |  |
| --- | --- | --- | --- | --- | --- | --- | --- | --- | --- | --- | --- | --- | --- |
|  |  | RRR | Low CI | High CI | p-value | RRR | Low CI | High CI | p-value | RRR | Low CI | High CI | p-value |
| **Female vs. male** |  | 0.85 | 0.51 | 1.40 | 0.519 | 0.75 | 0.53 | 1.07 | 0.110 | 0.74 | 0.51 | 1.06 | 0.097 |
| **Age (per 10-year increase)** |  | 1.16 | 1.01 | 1.35 | 0.042 | 1.21 | 1.08 | 1.35 | 0.001 | 0.94 | 0.84 | 1.04 | 0.240 |
| **Hispanic vs. non-Hispanic** |  | 0.95 | 0.43 | 2.08 | 0.893 | 1.34 | 0.83 | 2.15 | 0.230 | 1.29 | 0.76 | 2.17 | 0.347 |
| **Race** | White | 1.00 | - | - | 0.864 | 1.00 | - | - | 0.072 | 1.00 | - | - | <0.001 |
|  | Black/African American | 1.16 | 0.63 | 2.10 |  | 1.63 | 1.05 | 2.53 |  | 2.00 | 1.28 | 3.12 |  |
|  | Asian | 0.96 | 0.44 | 2.09 |  | 0.86 | 0.48 | 1.54 |  | 0.66 | 0.36 | 1.22 |  |
|  | Other | 0.62 | 0.14 | 2.75 |  | 0.78 | 0.29 | 2.08 |  | 1.37 | 0.53 | 3.55 |  |
| **Rural vs. urban** |  | 0.75 | 0.28 | 1.47 | 0.406 | 0.79 | 0.49 | 1.26 | 0.317 | 0.84 | 0.52 | 1.37 | 0.487 |
| **Political affiliation** | Democrat | 1.00 | - | - | 0.150 | 1.00 | - | - | <0.001 | 1.00 | - | - | 0.003 |
|  | Republican | 1.73 | 1.00 | 2.99 |  | 2.36 | 1.51 | 3.69 |  | 2.16 | 1.38 | 3.37 |  |
|  | Independent/ other | 1.27 | 0.68 | 2.35 |  | 1.20 | 0.80 | 1.81 |  | 1.22 | 0.79 | 1.87 |  |

\*The vaccine features group includes those who preferred a single dose vaccine, a two-dose vaccine or to vaccinate once only

**Table 3:** Choice experiment attributes and attribute levels.

| Attributes: | Attribute levels: |
| --- | --- |
| **Vaccination location** | Local pharmacy (e.g., Walgreens, CVS, other) |
|  | Health service (e.g., hospital, doctor office, community center) |
|  | Local pop-up vaccination service (e.g., fire station, community center, school, workplace) |
|  | At your home (door-to-door vaccination) |
|  | Large vaccination site supported by National Guard (e.g., convention center, stadium) |
| **Waiting time at vaccination site** | Immediate service |
|  | 1 hours |
|  | 2 hours |
| **Vaccination appointment scheduling** | Online website |
|  | Phone Call |
|  | No scheduling required (drop-in appointment) |
| **Number of doses required per vaccination episode** | 1 dose only |
|  | 2 doses one month apart |
| **Vaccination enforcement** | Vaccination is completely voluntary (not enforced in any way) |
|  | Vaccination is required for air travel (local and foreign) |
|  | Vaccination is required to attend work/school |
|  | Vaccination is required to attend public recreational spaces, e.g. movies, concerts, conferences) |
| **Who has already received the vaccine in your community?** | No one I know |
|  | A few people I know |
|  | Almost everyone I know |
| **Vaccine frequency** | One vaccination required for life-long protection against severe COVID-19 infection |
|  | Annual vaccinations required to maintain protection against severe COVID-19 infection |

**Fig 5:**
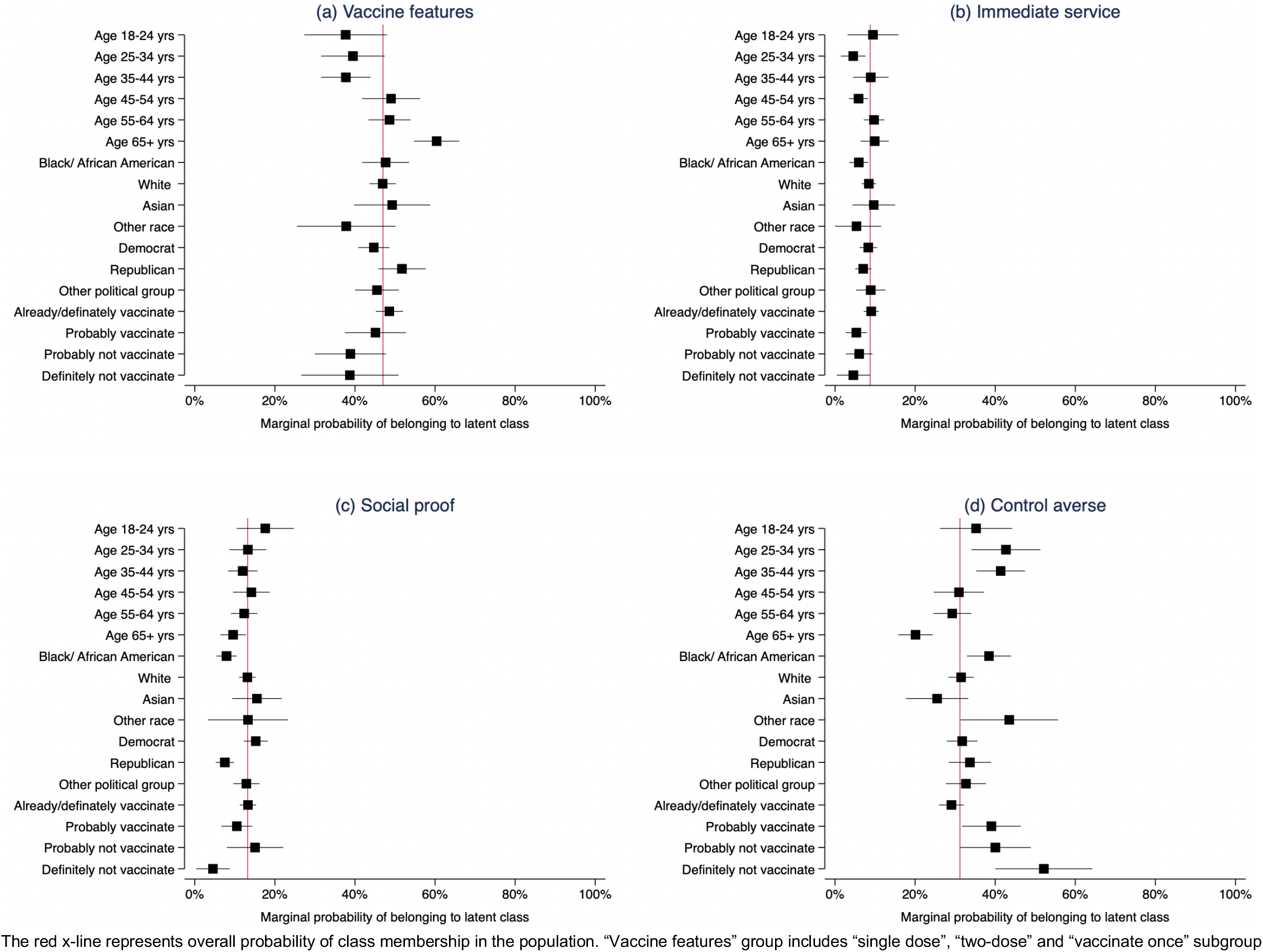
Marginal probability of belonging to latent class group by age, race, political affiliation and vaccination intention. The red x-line represents overall probability of class membership in the population. “Vaccine features” group includes “single dose”, “two-dose” and “vaccinate once” subgroup

## Discussion

This survey and choice experiment evaluating preferences of the US public for COVID-19 vaccine distribution campaign features conducted between March 15 to 22, 2021 demonstrated that US populations prefer to vaccinate under campaigns that provided single rather than multiple vaccine doses, where waiting times at vaccination sites are short and where vaccination is offered at health facilities, pharmacies or in homes and not at mass vaccination sites supported by the national guard. Additionally, the prospect of vaccinating once to get lifelong immunity as opposed to annual vaccination was highly appealing, with the strongest preferences across attributes seen for this potential future vaccine feature. Negative preferences were seen consistently for all vaccination enforcement strategies – air travel, work or school and recreational gatherings. The preferences of those who showed some vaccine hesitancy by stating that they would “probably get vaccinated”, “probably NOT get vaccinated” or “definitely NOT get vaccinated” indicated that vaccination enforcement would reduce their likelihood of vaccinating compared to voluntary vaccination. Latent class analyses revealed six preference subgroups in the population, three groups (approximately half the population) prioritized inherent “vaccine features” and preferred single dose or two dose vaccines, or to vaccinate once rather than annually. A small sub-group (8.8%), who were concerned about services at the vaccination site, indicated that they would be more likely to vaccinate if they could receive “immediate service”. A fifth “control averse” group (31% of the population) were mostly indifferent to vaccine and vaccine campaign features, but showed consistent negative preferences for all enforcement strategies. And the final “wait and see” group (13%) preferred to see others in their community vaccinated before vaccinating themselves. Compared to the “wait and see” group, those who were control averse were twice as likely to be republicans rather than democrats and be Black African/American rather than white.

These data provide support for the fundamental behavioral science principle that people are effort minimizers, frequently making health decisions based on the options that are easy and require the least effort (1, 17). Main preferences for reduced waiting time, single vaccine doses, vaccinating once and in-home vaccination highlight the importance of simplifying access and reducing the number of vaccinations required to achieve adequate levels of immunity. And although those who have already vaccinated or definitely intend to do so will likely vaccinate under any conditions, for those who continue to be “fence sitters” and deliberate - the “probably get vaccinated” or “probably NOT get vaccinated” groups - adjusting services to minimize “friction” in taking up vaccination could help facilitate action (18, 19). Restoring confidence in single dose vaccines, and streamlining vaccine services will be critical for improving uptake among unvaccinated and for future vaccination campaigns.

Minimizing effort may also extend to offering the public a choice of services, an approach that could promote autonomy and intrinsic motivation, giving some semblance of control back to individuals who, in recent times, have repeatedly been called on to make choices for the good of the ‘herd’. Self-determination theory proposes that providing choice enhances intrinsic motivation by promoting perceived agency and personal control over action and is more effective in settings where the group norm is individualism rather than collectivism (20, 21). Reports in the lay press of some “shopping around” or deferring vaccination to obtain preferred vaccine features are mirrored in our data (22, 23), with approximately half of the population more likely to vaccinate under scenarios where their strongly preferred vaccine or campaign features are available, specifically dosage schedules and vaccination frequency. Further preference drivers such as vaccine efficacy and side effect profiles also significantly influence choice (24-28). Although mass sites, at times supported by the national guard, have been highly effective in increasing vaccination rates across the US (29), evidence from this survey and those in other settings indicate that overwhelming people prefer health centers or pharmacies to community locations or mass vaccination sites (24, 26, 28). Even though the rationale for vaccine preferences may not always be based on scientific principles, driven by other belief systems, lay media reports or experiences – offering the public choices can hand some control back to individuals, a strategy also embraced by CDC vaccination campaigns aiming to appeal directly to individuals and promote autonomy with the campaign slogan - “It’s Up to You” (30). Given, the substantial heterogeneity of preferences, the increasing variety of vaccines becoming available, increasing vaccine supply and plateauing vaccine uptake in the US, developing simple and efficient systems that allow individuals to take more control and choose their preferred vaccine and vaccination strategy could improve vaccine uptake, a strategy which may not have been feasible or equitable during the early stages of the pandemic but may be more of a reality as focus shifts to those who remain unvaccinated under current vaccination campaigns (31).

The smaller “social proof” preference subgroup - 13% of the population - indicated that they would be more likely to take up vaccination with increasing vaccination coverage in the community and potentially enforcement – this group were less concerned about vaccine features or services, they appeared sensitive to external influencing factors and their preferences suggest some responsiveness to social influence and ‘social proof’ of vaccine safety with increasing vaccination coverage (32, 33). The “social proof” group were more likely to be Democrats, those who were young or white race and may represent those who will respond to normative vaccination campaign messages regarding vaccine safety and coverage, nudges, incentives and vaccine mandates (33).

Mean preferences in population and within subgroups however, indicated that vaccine enforcement may reduce the likelihood of vaccination for some - vaccine mandates have the potential to both increase vaccination rates but can also to promote anti-vaccine sentiment (34). Strengthening anti-vaccine sentiment as a result of enforcement is of particular concern in the US where control aversion and psychological reactance may be significant drivers of vaccine hesitancy. Our exploration of preferences for COVID-19 vaccination enforcement strategies including restrictions to air travel, work or school attendance and recreational gatherings indicated that across the population, enforcement was perceived negatively and for the majority would reduce vaccination compared to voluntary vaccination, with at trend of increasing hesitancy with increasing vaccine hesitancy, and seen to a greater extent among those who were young, Black/African American or Republican. Such negative responses to vaccine mandates, may be related to the crowding out of voluntary commitment, a phenomenon also identified in Germany – where groups that were liberal, had low trust in the government and were at low risk for severe COVID-19 illness showed more “control aversion” and were more resistant to vaccination enforcement than their counterparts (35). Such control aversion may operate through several mechanisms including the impact of the policy message of distrust of the population to be socially responsible, the removal of an individual’s need to deliberate over vaccination driving out moral convictions to be “prosocial”, inherent mistrust of the government and health systems leading to reduced compliance, and compromising personal autonomy driving “psychological reactance” – the motivation to regain a freedom after it has been lost or threatened which leads people to resist (35, 36). This resistance to vaccination enforcement is not surprising, in North American culture which endorses individualism, independence and personal freedom rather than an interdependent collective sensibility, reactance may play a significant role because individuals’ have expectations of control and choice (33, 36, 37). In the US, vaccine mandates may impinge on First amendment freedoms and have been found to increase anger among those already vaccine hesitant (38, 39). In Australia, in 2016 the government introduced legislation to reduce government payments to those who did not comply with childhood vaccinations, among non-vaccinating parents this resulted in the development of a greater commitment to their decision not to vaccinate and an increased willingness to become involved in protest action (40). The issue of governmental mistrust and affective political polarization which promotes distrust of opposing parties is another key feature of the current American narrative which may drive reluctance to vaccinate under enforcement, vaccine campaigns during the Ebola outbreak in Liberia 2019 were most effective where trust in the government was high, a scenario which may only apply to some populations in the US today (33). It is unclear to what extent these factors would impact COVID-19 vaccine uptake in the US, globally enforcement strategies have resulted in modest increases in childhood vaccination rates (34), and preferences elicited in choice experiments do not always translate to action, our findings however combined with relatively high rates of vaccine hesitancy in the US, suggest that enforcement strategies may be met with significant discontent, encouraging groups to double down on intentions not to vaccinate – a potential unintended consequence of vaccine mandates which could undermine efforts to expand vaccine coverage nationally.

These findings are strengthened by the use of a representative sample of the US population and the inclusion of preliminary survey questions which mirror the US Census Pulse survey allowing for re-weighting to match the US population distribution by age, gender, race, schooling and vaccination status and intention for the corresponding time period of our survey. Results of discrete choice experiments however, particularly in public health, must always be interpreted within a broader body of knowledge and understood within the boundaries of rational choice theory – which posits that individuals will make rationale choices to maximize their utility or happiness (41). Rational choice theory is strongly based on the assumptions which inform the model, namely the assumption of completeness – that an individual has a preference for one scenario over another or is indifferent but never completely uncertain, and the assumption of transitivity - that if attribute A is preferred to attribute B and attribute B is preferred to attribute C, attribute A must be preferred to attribute C. And although revealed preferences frequently match stated preferences, these assumptions may not always hold in real life decision making, and as a result utilities cannot always perfectly predict uptake of an intervention or service, particularly among those who show negative preferences (42). Results of willingness to pay or in this case willingness to wait analyses may also be limited by inherent assumptions - in this case the assumed linear relationship between waiting time and choice beyond the two hours presented in the experiment - in reality participants may not be willing to wait as long as 8 hours for a vaccination that affords life-long immunity rather than annual vaccination but such analyses can be a useful representation of the extent of the trade-offs that participants are willing to make to get the what they want and should be interpreted in this light. A further limitation of this study was the use of an online sample which may have influenced preferences for online vaccination appointment scheduling.

Facilitating ease of vaccination by offering a choice of vaccine brands, vaccination locations and promoting voluntary vaccination, may be best aligned with public preferences for COVID-19 vaccination campaigns features. Several strategies to simplify vaccination processes are already being implemented by various state health departments - our nationally representative data support their use more broadly across the US. Our data further emphasize the potential risks of vaccination enforcement strategies, which could potentially undermine vaccination efforts in an individualistic society such as the US, where trust in the government is low and where vaccine hesitancy is closely associated with political alliances.

## Data Availability

On publication the data will be made available in supplementary materials for the journal

## References

1. Volpp KG, Loewenstein G, Buttenheim AM. Behaviorally Informed Strategies for a National COVID-19 Vaccine Promotion Program. JAMA. 2021;325(2):125–6.

2. Bridges JF, Hauber AB, Marshall D, Lloyd A, Prosser LA, Regier DA, et al. Conjoint analysis applications in health--a checklist: a report of the ISPOR Good Research Practices for Conjoint Analysis Task Force. Value in health : the journal of the International Society for Pharmacoeconomics and Outcomes Research. 2011;14(4):403–13.

3. Reed Johnson F, Lancsar E, Marshall D, Kilambi V, Muhlbacher A, Regier DA, et al. Constructing experimental designs for discrete-choice experiments: report of the ISPOR Conjoint Analysis Experimental Design Good Research Practices Task Force. Value in health : the journal of the International Society for Pharmacoeconomics and Outcomes Research. 2013;16(1):3–13.

4. Helter TM, Boehler CEH. Developing attributes for discrete choice experiments in health: a systematic literature review and case study of alcohol misuse interventions. Journal of substance use. 2016;21(6):662–8.

5. Obadha M, Barasa E, Kazungu J, Abiiro GA, Chuma J. Attribute development and level selection for a discrete choice experiment to elicit the preferences of health care providers for capitation payment mechanism in Kenya. Health Economics Review. 2019;9(1):30.

6. Sawtooth Software [Available from: https://www.sawtoothsoftware.com/.

7. Johnson FR, Yang JC, Reed SD. The Internal Validity of Discrete Choice Experiment Data: A Testing Tool for Quantitative Assessments. Value in health : the journal of the International Society for Pharmacoeconomics and Outcomes Research. 2019;22(2):157–60.

8. Orme B. Getting Started with Conjoint Analysis: Strategies for Product Design and Pricing Research. Second Edition ed. Madison, Wisconsin, USA: Research Publishers LLC; 2010.

9. United States Census Bureau. Household Pulse Survey: Measuring Household Experiences during the Coronavirus Pandemic 29 March 2021 [Available from: https://www.census.gov/data/experimental-data-products/household-pulse-survey.html.

10. Qualtrics. 2005 [Available from: https://www.qualtrics.com.

11. Lancsar E, Louviere J. Conducting discrete choice experiments to inform healthcare decision making: a user’s guide. Pharmacoeconomics. 2008;26(8):661–77.

12. Campbell D, Erdem S. Including Opt-Out Options in Discrete Choice Experiments: Issues to Consider. The patient. 2019;12(1):1–14.

13. Veldwijk J, Lambooij MS, de Bekker-Grob EW, Smit HA, de Wit GA. The effect of including an opt-out option in discrete choice experiments. PloS one. 2014;9(11):e111805–e.

14. Daly A, Dekker T, Hess S. Dummy coding vs effects coding for categorical variables: Clarifications and extensions. Journal of Choice Modelling. 2016;21:36–41.

15. Hole AR. Fitting mixed logit models by using maximum simulated likelihood. Stata Journal. 2007;7(3):388–401.

16. Mori M, Krumholz HM, Allore HG. Using Latent Class Analysis to Identify Hidden Clinical Phenotypes. JAMA. 2020;324(7):700–1.

17. Hallsworth M. Rethinking public health using behavioural science. Nature human behaviour. 2017;1(9):612.

18. Service O, Hallsworth M, Halperin D, Algate F, Gallagher R, Nguyen S, et al. EAST: Four simple ways to apply behavioural insights 2014 [Available from: https://www.bi.team/publications/east-four-simple-ways-to-apply-behavioural-insights/.

19. Betsch C, Korn L, Holtmann C. Don’t try to convert the antivaccinators, instead target the fence-sitters. Proceedings of the National Academy of Sciences. 2015;112(49):E6725.

20. Hagger MS, Rentzelas P, Chatzisarantis NLD. Effects of individualist and collectivist group norms and choice on intrinsic motivation. Motivation and Emotion. 2014;38(2):215–23.

21. Deci EL, Ryan RM. The “What” and “Why” of Goal Pursuits: Human Needs and the Self-Determination of Behavior. Psychological Inquiry. 2000;11(4):227–68.

22. Yankowski P, Cuda A. Shopping around for a COVID-19 vaccine? Some say it’s not worth it. Connecticut Post. 2021 March 19, 2021.

23. Sotnick K. Health Officials Say People Should Not ‘Shop Around’ for Preferred Vaccine. NBC Boston. 2021.

24. Craig BM. United States COVID-19 Vaccination Preferences (CVP): 2020 Hindsight. The patient. 2021;14(3):309–18.

25. Kreps S, Prasad S, Brownstein JS, Hswen Y, Garibaldi BT, Zhang B, et al. Factors Associated With US Adults’ Likelihood of Accepting COVID-19 Vaccination. JAMA Network Open. 2020;3(10):e2025594–e.

26. McPhedran R, Toombs B. Efficacy or delivery? An online Discrete Choice Experiment to explore preferences for COVID-19 vaccines in the UK. Economics Letters. 2021;200:109747.

27. Motta M. Can a COVID-19 vaccine live up to Americans’ expectations? A conjoint analysis of how vaccine characteristics influence vaccination intentions. Social Science & Medicine. 2021;272:113642.

28. Schwarzinger M, Watson V, Arwidson P, Alla F, Luchini S. COVID-19 vaccine hesitancy in a representative working-age population in France: a survey experiment based on vaccine characteristics. The Lancet Public Health. 2021;6(4):e210–e21.

29. Goralnick E, Kaufmann C, Gawande AA. Mass-Vaccination Sites — An Essential Innovation to Curb the Covid-19 Pandemic. New England Journal of Medicine. 2021.

30. Ad Council. The Ad Council and COVID Collaborative Reveal ‘It’s Up To You’ Campaigns to Educate Millions of Americans about COVID-19 Vaccines 2021 [Available from: https://www.adcouncil.org/press-releases/the-ad-council-and-covid-collaborative-reveal-its-up-to-you-campaigns-to-educate-millions-of-americans-about-covid-19-vaccines.

31. Kramer DB, Opel DJ, Parasidis E, Mello MM. Choices in a Crisis — Individual Preferences among SARS-CoV-2 Vaccines. New England Journal of Medicine. 2021;384(17):e62.

32. Hallsworth M, Mirupi S, Toth C. 2021 Mar 15, 2021. [cited 2021 Apr 26, 2021]. Available from: https://www.bi.team/blogs/four-messages-that-can-increase-uptake-of-the-covid-19-vaccines/.

33. Bavel JJV, Baicker K, Boggio PS, Capraro V, Cichocka A, Cikara M, et al. Using social and behavioural science to support COVID-19 pandemic response. Nature Human Behaviour. 2020;4(5):460–71.

34. Omer SB, Betsch C, Leask J. Mandate vaccination with care. Nature. 2019;571(7766):469–72.

35. Schmelz K. Enforcement may crowd out voluntary support for COVID-19 policies, especially where trust in government is weak and in a liberal society. Proceedings of the National Academy of Sciences. 2021;118(1):e2016385118.

36. Steindl C, Jonas E, Sittenthaler S, Traut-Mattausch E, Greenberg J. Understanding Psychological Reactance: New Developments and Findings. Z Psychol. 2015;223(4):205–14.

37. Miron AM, Brehm JW. Reactance Theory - 40 Years Later. Zeitschrift für Sozialpsychologie. 2006;37(1):9–18.

38. Gostin LO, Cohen IG, Shaw J. Digital Health Passes in the Age of COVID-19: Are “Vaccine Passports” Lawful and Ethical? JAMA. 2021.

39. Betsch C, Böhm R. Detrimental effects of introducing partial compulsory vaccination: experimental evidence. Eur J Public Health. 2016;26(3):378–81.

40. Helps C, Leask J, Barclay L. “It just forces hardship”: impacts of government financial penalties on non-vaccinating parents. Journal of public health policy. 2018;39(2):156–69.

41. Lancaster KJ. A New Approach to Consumer Theory. Journal of Political Economy. 1966;74(2):132–57.

42. de Bekker-Grob Ew, Donkers B, Bliemer MCJ, Veldwijk J, Swait JD. Can healthcare choice be predicted using stated preference data? Social Science & Medicine. 2020;246:112736.

