## Supplementary materials for "Strategies that make vaccination easy and promote autonomy could increase COVID-19 vaccination in those who remain hesitant"

Appendix S1: Survey tool

### Default Question Block

The COVID-19 Vaccine Distribution Survey

We invite you to participate in a research study being conducted by investigators from Washington University in St. Louis. This survey will help us determine how best to distribute COVID-19 vaccines in your community. The survey will take approximately 15-20 minutes to complete. You can complete the survey on your phone, however the best visibility will be on a tablet or computer.

Your participation is voluntary and anonymous. By completing this survey you will be contributing to a better understanding of how best to distribute COVID-19 vaccines in your community. There are no foreseeable risks to your participation, however you should note that the survey and storage are not hosted by or at Washington University, the data will hosted by third party survey administration companies - Qualtrics and Sawtooth Software. The Washington University information Security Office have however reviewed and approved of the information security of these survey companies. You will be compensated for your participation according to your agreement with the Qualtrics survey team.

The first portion of the survey will ask questions about your demographic characteristics, employment and COVID-19 experiences. The second portion of the survey will ask you to compare two COVID-19 vaccination scenarios across 10 questions and select the option you prefer the most.

If you have questions for the research team, please contact Dr. Ingrid Eshun-Wilson, or you may contact the Human Research Protection Office at 1-800-438-0445

What is your current age in years?

What is your gender?

- Male
- Female
- Other
- Prefer not to answer

Are you Hispanic, Latino or Spanish origin

- No, not of Hispanic, Latino, or Spanish origin
- Yes, Mexican, Mexican American, Chicano
- Yes, Puerto Rican
- Yes, Cuban
- Yes, another Hispanic, Latino, or Spanish origin

What is your race?

- White
- Black or African American
- American Indian or Alaska Native

- Native Hawaiian
- Asian Indian
- Chinese
- Filipino
- Japanese
- Korean
- Vietnamese
- Other Asian
- Samoan
- Other Pacific Islander
- Prefer not to answer

What is the highest degree or level of school you have completed?

- Less than high school
- Some high school
- High school graduate or equivalent (for example GED)
- Some college, but degree not received or is in progress
- Associate's degree (for example, AA, AS)
- Bachelor's degree (for example BA, BS, AB)
- Graduate degree (for example master's, professional, doctorate)
- Prefer not to answer

What is your political affiliation?

- Democrat
- Republican
- Other
- Unaffiliated
- Prefer not to answer

What is your zip code?

You can leave the zip code blank if you prefer us not to collect this information.

Do you have any children?

- ◯ Yes
- ◯ No

How many children do you have?

What are their ages? (Select all that apply)

- ☐ 0 to 5 years
- ☐ 6 to 13 years
- ☐ 14 to 18 years
- ☐ 19 to 25 years
- ☐ 26 years or older

Have you, or has anyone in your household experienced a loss of employment income since March 13, 2020?

- ◯ Yes
- ◯ No

Do you expect that you or anyone in your household will experience a loss of employment income in the next 4 weeks because of the coronavirus pandemic?

- Yes
- No

Now we are going to ask about your employment.

In the last 7 days, did you do ANY work for either pay or profit?

- Yes
- No

What is your main reason for not working for pay or profit?
I did not work because:

- I did not want to be employed at this time
- I am/was sick with coronavirus symptoms
- I am/was caring for someone with coronavirus symptoms
- I am/was caring for children not in school or daycare
- I am/was caring for an elderly person
- I was concerned about getting or spreading the coronavirus
- I am/was sick (not coronavirus related) or disabled
- I am retired
- My employer experienced a reduction in business (including furlough) due to coronavirus pandemic
- I am/was laid off due to coronavirus pandemic
- My employer closed temporarily due to the coronavirus pandemic
- My employer went out of business due to the coronavirus pandemic
- Other reason

Since March 13, 2020 have you applied for Unemployment Insurance (UI) benefits?

- Yes
- No

Since March 13, 2020, did you receive Unemployment Insurance (UI) benefits?

- Yes
- No

Now we are going to ask about your experiences and beliefs about COVID-19 vaccination and infection.

Has a COVID-19 vaccine been made available to you yet? (Are you eligible for a vaccine?)

- Yes
- No
- Not sure

Have you receive some or all the required COVID-19 vaccine doses?

- Yes
- No

If a vaccine to prevent COVID-19 is available to you, would you…

- Definitely get a vaccine

- ○ Probably get a vaccine
- ○ Probably NOT get a vaccine
- ○ Definitely NOT get a vaccine

Which of the following, if any, are reasons that you would NOT get vaccinated? (Select all that apply)

- ☐ I am concerned about possible side effects of a COVID-19 vaccine
- ☐ I don't know if a COVID-19 vaccine will work
- ☐ I don't believe I need a COVID-19 vaccine
- ☐ I don't like vaccines
- ☐ My doctor has not recommended it
- ☐ I plan to wait and see if it is safe and may get it later
- ☐ I think other people need it more than I do right now
- ☐ I am concerned about the cost of a COVID-19 vaccine
- ☐ I don't trust COVID-19 vaccines
- ☐ I don't trust the government
- ☐ Other reason

Why do you believe that you don't need a COVID-19 vaccine? (Select all that apply)

- ☐ I already had COVID-19
- ☐ I am not a member of a high-risk group
- ☐ I plan to use masks or other precautions instead
- ☐ I don't believe COVID-19 is a serious illness
- ☐ I don't think vaccines are beneficial
- ☐ Other reason

What is the reason you have not received any COVID-19 vaccination doses yet?

- I tried but could not get an appointment
- The vaccination site is too far away (transport is difficult)
- I could not find information on where to get a vaccination
- Other reason

Has a doctor or other health care provider ever told you that you have COVID-19?

- Yes
- No
- Not sure

In the next section you will be asked choice survey questions. A choice survey is a type of survey where you are presented with a few scenarios (e.g. scenario A and scenario B), in each scenario the features of a service or product are changed slightly. After you review the features of each scenario you determine which one you prefer and you make a selection based on which you think you like more. In this simple example below you would choose the ice cream you prefer.

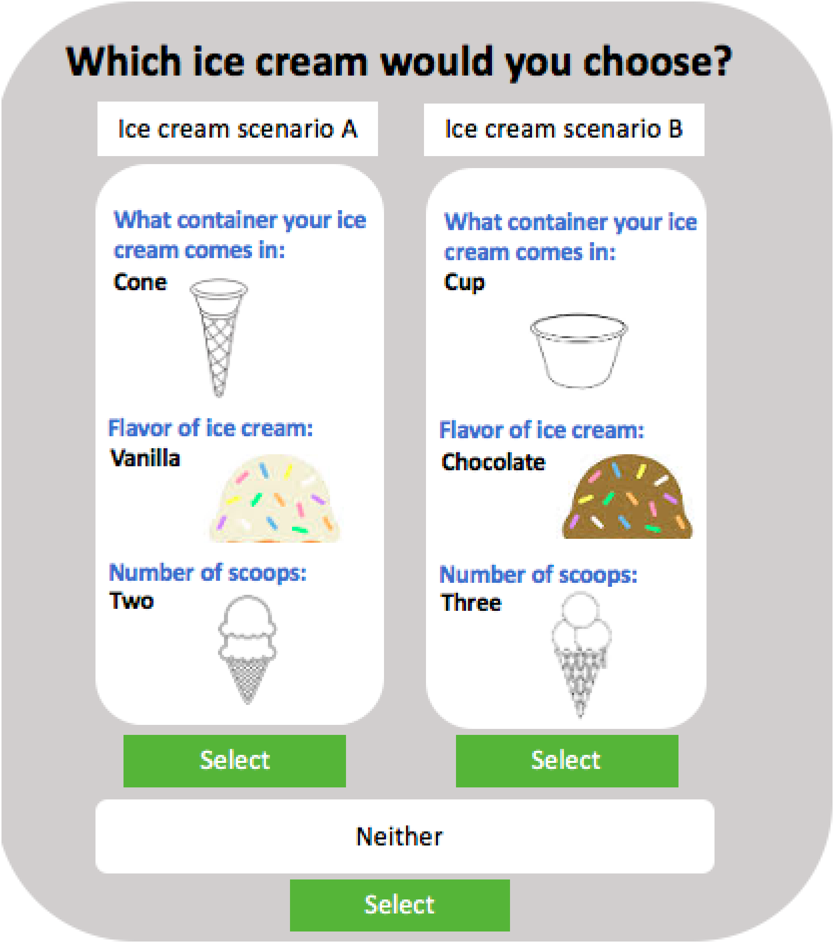


In the following 10 choice survey questions you will be asked to choose between two COVID-19 vaccination scenarios (situations) similar to the ice cream example above, please review these vaccination scenarios carefully and decide under which scenario you would be more likely to vaccinate for COVID-19.

You can assume that for each scenario the COVID-19 vaccine is 90% effective in preventing severe COVID-19 infection (hospitalization or death) and that serious adverse events are rare, occurring in less than 1 in 1,000,000 people.

Please complete all 10 questions although they may appear repetitive, your thoughtful consideration of which COVID-19 vaccination distribution features you prefer could help with the design of vaccination programs.

You are not eligible for this survey. Thank you.

CBC_Random1

After reviewing the seven features of vaccination scenario A and comparing these with the seven features of vaccination scenario B, please select the vaccination scenario you would prefer for COVID-19 vaccination? If you would not vaccinate under either of these two vaccine scenarios, select the third 'neither' option.

(1 of 10)

**COVID-19 vaccine scenario A**

**How long you wait at the site to get your vaccination:**

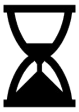

**1 hour**

**How many people in your community have already received the vaccine:**

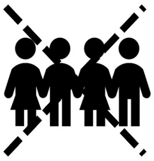

**No one you know**

**How many vaccine doses you need per vaccination:**

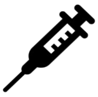

**1 Dose**

**One dose**
(one injection for full vaccination)

**How you can schedule your appointment:**

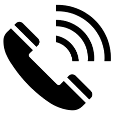

**Book through phone call**

**How frequently you need to vaccinate:**

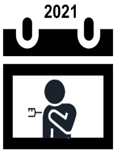


**One vaccination episode only**
(protection for more than 10 years)

**COVID-19 vaccine scenario B**

**How long you wait at the site to get your vaccination:**

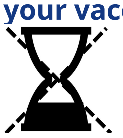

**Immediate service**

**How many people in your community have already received the vaccine:**

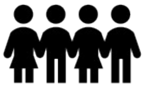

**A few people you know**

**How many vaccine doses you need per vaccination:**

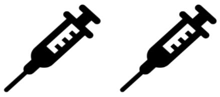

**2 Doses**

**Two doses**
(two injections one month apart)

**How you can schedule your appointment:**

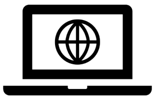

**Book online**

**How frequently you need to vaccinate:**

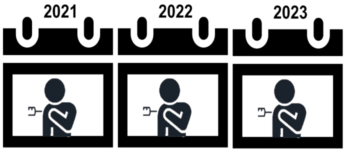


**Annual vaccination**
(vaccination needed yearly, similar to flu)

**Where you can get your vaccination:**

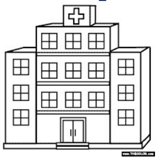

**Health service**
(e.g. hospital, doctor office, community center)

**How vaccination is enforced:**

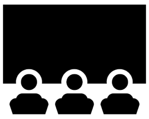

**Vaccination is required to attend public recreational spaces**
(e.g. movies, concerts, conferences)

CBC_Random1 Select

**Where you can get your vaccination:**

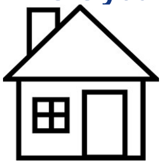

**At your home**
(door to door vaccination)

**How vaccination is enforced:**

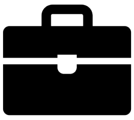

**Vaccination is required to attend work/college**

CBC_Random1 Select

**Neither scenario A nor scenario B**

NONE: I wouldn't choose any of these.

CBC_Random1

Select

Back Next

0% 100%

Table S1: Weighted mean preferences – total population

| Attribute | Relative utilities | | | | Standard deviation | | | |
|---|---|---|---|---|---|---|---|---|
| | Utility | Low CI | High CI | p-value | SD | Low CI | High CI | p-value |
| Opt-out | -1.77 | -1.96 | -1.58 | <0.001 | 0.50 | 0.21 | 0.79 | 0.001 |
| Vaccinate at pharmacy vs. health center | 0.07 | -0.04 | 0.19 | 0.222 | 0.65 | 0.39 | 0.90 | <0.001 |
| Vaccinate at community venue vs. health center | -0.13 | -0.24 | -0.02 | 0.022 | 0.93 | 0.74 | 1.13 | <0.001 |
| Vaccinate at home vs. health center | -0.10 | -0.27 | 0.07 | 0.239 | 0.69 | 0.51 | 0.87 | <0.001 |
| Vaccinate at mass site vs. health center | -0.29 | -0.39 | -0.18 | <0.001 | -0.24 | -0.64 | 0.15 | 0.221 |
| Wait for 1 hr vs. immediate service | -0.21 | -0.32 | -0.11 | <0.001 | 0.53 | 0.35 | 0.71 | <0.001 |
| Wait for 2 hrs vs. immediate service | -0.54 | -0.64 | -0.43 | <0.001 | 0.53 | 0.37 | 0.70 | <0.001 |
| Phone vs. online appointment booking | -0.06 | -0.15 | 0.02 | 0.145 | 0.41 | 0.18 | 0.63 | <0.001 |
| Drop in (no booking) vs. online appointment booking | 0.05 | -0.03 | 0.13 | 0.251 | 1.07 | 0.97 | 1.18 | <0.001 |
| Vaccinate annually vs. once | -0.79 | -0.89 | -0.70 | <0.001 | 0.23 | -0.13 | 0.60 | 0.209 |
| Enforcement for air travel vs. no enforcement | -0.13 | -0.23 | -0.04 | 0.006 | -0.40 | -0.64 | -0.17 | 0.001 |
| Enforcement for work/school vs. no enforcement | -0.20 | -0.30 | -0.10 | <0.001 | -0.22 | -0.50 | 0.06 | 0.126 |
| Enforcement for recreation vs. no enforcement | -0.05 | -0.15 | 0.05 | 0.298 | 0.36 | 0.06 | 0.65 | 0.017 |
| A few in the community vaccinated vs. no one | 0.36 | 0.28 | 0.43 | <0.001 | -0.49 | -0.66 | -0.32 | <0.001 |
| Almost everyone in the community vaccinated vs. no one | 0.48 | 0.39 | 0.56 | <0.001 | 0.97 | 0.83 | 1.10 | <0.001 |
| Two vaccine doses vs. a single dose | -0.29 | -0.37 | -0.20 | <0.001 | 0.50 | 0.21 | 0.79 | 0.001 |

Table S2(a): Weighted mean preferences – Already vaccinated

| Attribute | Relative utilities | | | | Standard deviation | | | |
|---|---|---|---|---|---|---|---|---|
| | Utility | Low CI | High CI | p-value | SD | Low CI | High CI | p-value |
| Opt-out | -1.79 | -2.10 | -1.49 | <0.001 | | | | |
| Vaccinate at pharmacy vs. health center | 0.17 | -0.03 | 0.37 | 0.089 | -0.30 | -1.02 | 0.43 | 0.422 |
| Vaccinate at community venue vs. health center | -0.09 | -0.27 | 0.08 | 0.300 | 0.72 | 0.36 | 1.07 | <0.001 |
| Vaccinate at home vs. health center | -0.01 | -0.29 | 0.27 | 0.939 | 1.08 | 0.78 | 1.37 | <0.001 |
| Vaccinate at mass site vs. health center | -0.31 | -0.48 | -0.14 | <0.001 | 0.69 | 0.41 | 0.97 | <0.001 |
| Wait for 1 hr vs. immediate service | -0.20 | -0.37 | -0.02 | 0.027 | 0.39 | 0.07 | 0.72 | 0.017 |
| Wait for 2 hrs vs. immediate service | -0.65 | -0.84 | -0.47 | <0.001 | 0.82 | 0.44 | 1.20 | <0.001 |
| Phone vs. online appointment booking | -0.09 | -0.23 | 0.05 | 0.201 | 0.56 | 0.28 | 0.84 | <0.001 |
| Drop in (no booking) vs. online appointment booking | 0.14 | 0.00 | 0.27 | 0.044 | 0.46 | 0.13 | 0.79 | 0.007 |
| Vaccinate annually vs. once | -0.96 | -1.13 | -0.80 | <0.001 | 1.10 | 0.95 | 1.25 | <0.001 |
| Enforcement for air travel vs. no enforcement | 0.01 | -0.14 | 0.17 | 0.879 | -0.27 | -0.89 | 0.35 | 0.399 |
| Enforcement for work/school vs. no enforcement | -0.14 | -0.31 | 0.03 | 0.110 | -0.38 | -0.69 | -0.07 | 0.015 |
| Enforcement for recreation vs. no enforcement | 0.10 | -0.05 | 0.25 | 0.211 | -0.07 | -0.56 | 0.41 | 0.769 |
| A few in the community vaccinated vs. no one | 0.45 | 0.34 | 0.57 | <0.001 | 0.16 | -0.06 | 0.37 | 0.152 |
| Almost everyone in the community vaccinated vs. no one | 0.57 | 0.44 | 0.70 | <0.001 | -0.42 | -0.75 | -0.08 | 0.015 |
| Two vaccine doses vs. a single dose | -0.13 | -0.30 | 0.03 | 0.112 | 1.10 | 0.83 | 1.38 | <0.001 |

Table S2(b): Weighted mean preferences – Definitely get vaccinated

| Attribute | Relative utilities | | | | Standard deviation | | | |
|---|---|---|---|---|---|---|---|---|
| | Utility | Low CI | High CI | p-value | SD | Low CI | High CI | p-value |
| Opt-out | -2.34 | -2.67 | -2.02 | <0.001 | | | | |
| Vaccinate at pharmacy vs. health center | -0.01 | -0.21 | 0.20 | 0.935 | 0.73 | 0.32 | 1.14 | <0.001 |
| Vaccinate at community venue vs. health center | -0.08 | -0.27 | 0.11 | 0.402 | 0.60 | 0.17 | 1.03 | 0.006 |
| Vaccinate at home vs. health center | -0.05 | -0.32 | 0.22 | 0.728 | 0.84 | 0.48 | 1.20 | <0.001 |
| Vaccinate at mass site vs. health center | -0.49 | -0.71 | -0.27 | <0.001 | 0.92 | 0.69 | 1.16 | <0.001 |
| Wait for 1 hr vs. immediate service | -0.28 | -0.46 | -0.10 | 0.003 | -0.03 | -0.51 | 0.46 | 0.912 |
| Wait for 2 hrs vs. immediate service | -0.58 | -0.78 | -0.37 | <0.001 | 0.49 | 0.15 | 0.83 | 0.004 |
| Phone vs. online appointment booking | 0.01 | -0.14 | 0.15 | 0.924 | -0.36 | -0.70 | -0.02 | 0.037 |
| Drop in (no booking) vs. online appointment booking | 0.07 | -0.09 | 0.23 | 0.370 | 0.54 | 0.22 | 0.86 | 0.001 |
| Vaccinate annually vs. once | -0.69 | -0.85 | -0.53 | <0.001 | 1.05 | 0.88 | 1.23 | <0.001 |
| Enforcement for air travel vs. no enforcement | -0.04 | -0.20 | 0.13 | 0.654 | 0.18 | -1.30 | 1.66 | 0.813 |
| Enforcement for work/school vs. no enforcement | -0.04 | -0.22 | 0.14 | 0.671 | 0.48 | -0.35 | 1.31 | 0.256 |
| Enforcement for recreation vs. no enforcement | 0.11 | -0.08 | 0.29 | 0.251 | 0.07 | -0.47 | 0.62 | 0.787 |
| A few in the community vaccinated vs. no one | 0.43 | 0.28 | 0.59 | <0.001 | 0.31 | -0.25 | 0.88 | 0.280 |
| Almost everyone in the community vaccinated vs. no one | 0.50 | 0.33 | 0.68 | <0.001 | 0.57 | 0.17 | 0.98 | 0.006 |
| Two vaccine doses vs. a single dose | -0.41 | -0.55 | -0.27 | <0.001 | 0.76 | 0.51 | 1.01 | <0.001 |

Table S2(c): Weighted mean preferences – Probably get vaccinated

| Attribute | Relative utilities | | | | Standard deviation | | | |
|---|---|---|---|---|---|---|---|---|
| | Utility | Low CI | High CI | p-value | SD | Low CI | High CI | p-value |
| Opt-out | -1.79 | -2.29 | -1.29 | <0.001 | | | | |
| Vaccinate at pharmacy vs. health center | 0.06 | -0.18 | 0.30 | 0.614 | -0.61 | -0.96 | -0.27 | 0.001 |
| Vaccinate at community venue vs. health center | -0.18 | -0.45 | 0.09 | 0.201 | 0.93 | 0.57 | 1.30 | <0.001 |
| Vaccinate at home vs. health center | -0.13 | -0.56 | 0.30 | 0.557 | 1.15 | 0.63 | 1.66 | <0.001 |
| Vaccinate at mass site vs. health center | -0.20 | -0.45 | 0.06 | 0.133 | 0.66 | 0.06 | 1.25 | 0.030 |
| Wait for 1 hr vs. immediate service | -0.17 | -0.42 | 0.08 | 0.185 | -0.37 | -0.63 | -0.11 | 0.005 |
| Wait for 2 hrs vs. immediate service | -0.23 | -0.49 | 0.03 | 0.078 | -0.41 | -0.80 | -0.02 | 0.041 |
| Phone vs. online appointment booking | -0.11 | -0.34 | 0.12 | 0.361 | -0.52 | -0.79 | -0.25 | <0.001 |
| Drop in (no booking) vs. online appointment booking | -0.06 | -0.27 | 0.15 | 0.587 | -0.17 | -0.48 | 0.15 | 0.300 |
| Vaccinate annually vs. once | -0.58 | -0.81 | -0.36 | <0.001 | 1.10 | 0.83 | 1.37 | <0.001 |
| Enforcement for air travel vs. no enforcement | -0.43 | -0.68 | -0.19 | <0.001 | -0.65 | -1.03 | -0.28 | 0.001 |
| Enforcement for work/school vs. no enforcement | -0.33 | -0.56 | -0.09 | 0.007 | 0.78 | 0.36 | 1.21 | <0.001 |
| Enforcement for recreation vs. no enforcement | -0.41 | -0.67 | -0.16 | 0.002 | 0.25 | -0.28 | 0.78 | 0.361 |
| A few in the community vaccinated vs. no one | 0.25 | 0.07 | 0.43 | 0.006 | 0.27 | -0.27 | 0.80 | 0.324 |
| Almost everyone in the community vaccinated vs. no one | 0.39 | 0.18 | 0.60 | <0.001 | -0.10 | -0.67 | 0.47 | 0.729 |
| Two vaccine doses vs. a single dose | -0.43 | -0.63 | -0.22 | <0.001 | 1.12 | 0.87 | 1.37 | <0.001 |

Table S2(d): Weighted mean preferences – Probably NOT get vaccinated

| Attribute | Relative utilities | | | | Standard deviation | | | |
|---|---|---|---|---|---|---|---|---|
| | Utility | Low CI | High CI | p-value | SD | Low CI | High CI | p-value |
| Opt-out | -1.68 | -2.20 | -1.15 | <0.001 | | | | |
| Vaccinate at pharmacy vs. health center | -0.09 | -0.42 | 0.24 | 0.580 | 0.12 | -0.41 | 0.66 | 0.655 |
| Vaccinate at community venue vs. health center | -0.40 | -0.87 | 0.07 | 0.096 | 1.16 | 0.22 | 2.09 | 0.016 |
| Vaccinate at home vs. health center | -0.64 | -1.08 | -0.21 | 0.004 | 1.27 | 0.79 | 1.75 | <0.001 |
| Vaccinate at mass site vs. health center | -0.34 | -0.75 | 0.07 | 0.100 | -0.91 | -1.54 | -0.27 | 0.005 |
| Wait for 1 hr vs. immediate service | -0.55 | -0.90 | -0.20 | 0.002 | -0.71 | -1.24 | -0.18 | 0.009 |
| Wait for 2 hrs vs. immediate service | -0.78 | -1.12 | -0.44 | <0.001 | 0.48 | -0.13 | 1.10 | 0.126 |
| Phone vs. online appointment booking | -0.08 | -0.40 | 0.24 | 0.611 | 0.56 | -0.06 | 1.18 | 0.078 |
| Drop in (no booking) vs. online appointment booking | 0.05 | -0.24 | 0.34 | 0.742 | 0.48 | -0.38 | 1.35 | 0.274 |
| Vaccinate annually vs. once | -0.70 | -0.92 | -0.48 | <0.001 | 0.86 | 0.64 | 1.09 | <0.001 |
| Enforcement for air travel vs. no enforcement | -0.64 | -0.93 | -0.34 | <0.001 | -0.55 | -0.93 | -0.17 | 0.005 |
| Enforcement for work/school vs. no enforcement | -0.97 | -1.39 | -0.56 | <0.001 | -0.79 | -1.33 | -0.25 | 0.004 |
| Enforcement for recreation vs. no enforcement | -0.77 | -1.08 | -0.45 | <0.001 | 0.38 | 0.02 | 0.73 | 0.038 |
| A few in the community vaccinated vs. no one | 0.39 | 0.10 | 0.67 | 0.008 | 0.42 | -0.17 | 1.01 | 0.164 |
| Almost everyone in the community vaccinated vs. no one | 0.58 | 0.29 | 0.86 | <0.001 | 0.40 | -0.22 | 1.02 | 0.201 |
| Two vaccine doses vs. a single dose | -0.32 | -0.57 | -0.06 | 0.014 | 1.08 | 0.32 | 1.83 | 0.005 |

Table S2(e): Weighted mean preferences – Definitely NOT get vaccinated

| Attribute | Relative utilities | | | | Standard deviation | | | |
|---|---|---|---|---|---|---|---|---|
| | Utility | Low CI | High CI | p-value | SD | Low CI | High CI | p-value |
| Opt-out | -1.57 | -2.51 | -0.63 | 0.001 | | | | |
| Vaccinate at pharmacy vs. health center | 0.00 | -0.49 | 0.49 | 0.999 | 1.03 | 0.28 | 1.78 | 0.007 |
| Vaccinate at community venue vs. health center | -0.11 | -0.58 | 0.37 | 0.651 | -0.81 | -2.15 | 0.52 | 0.234 |
| Vaccinate at home vs. health center | -0.02 | -0.80 | 0.76 | 0.957 | -1.38 | -1.94 | -0.81 | <0.001 |
| Vaccinate at mass site vs. health center | 0.20 | -0.27 | 0.67 | 0.399 | -0.22 | -0.91 | 0.47 | 0.527 |
| Wait for 1 hr vs. immediate service | -0.26 | -0.76 | 0.23 | 0.294 | 0.82 | 0.25 | 1.39 | 0.005 |
| Wait for 2 hrs vs. immediate service | -0.37 | -0.93 | 0.18 | 0.186 | 1.16 | 0.64 | 1.69 | <0.001 |
| Phone vs. online appointment booking | -0.09 | -0.53 | 0.35 | 0.696 | -0.72 | -1.35 | -0.09 | 0.025 |
| Drop in (no booking) vs. online appointment booking | -0.12 | -0.63 | 0.40 | 0.657 | 0.48 | -0.13 | 1.09 | 0.121 |
| Vaccinate annually vs. once | -0.44 | -0.86 | -0.01 | 0.047 | 0.83 | 0.34 | 1.31 | 0.001 |
| Enforcement for air travel vs. no enforcement | -0.96 | -1.52 | -0.39 | 0.001 | 1.20 | 0.70 | 1.71 | <0.001 |
| Enforcement for work/school vs. no enforcement | -0.87 | -1.50 | -0.24 | 0.007 | 1.07 | 0.29 | 1.84 | 0.007 |
| Enforcement for recreation vs. no enforcement | -1.01 | -1.48 | -0.54 | <0.001 | -0.62 | -1.30 | 0.06 | 0.076 |
| A few in the community vaccinated vs. no one | -0.07 | -0.41 | 0.27 | 0.681 | 0.40 | -0.14 | 0.94 | 0.149 |
| Almost everyone in the community vaccinated vs. no one | 0.33 | -0.08 | 0.74 | 0.113 | -0.54 | -1.24 | 0.15 | 0.126 |
| Two vaccine doses vs. a single dose | -0.38 | -0.70 | -0.05 | 0.022 | 0.86 | 0.48 | 1.24 | <0.001 |

Table S3(a): Weighted mean preferences – "single dose" latent class group

| Attribute | Relative utilities | | | | Standard deviation | | | |
|---|---|---|---|---|---|---|---|---|
| | Utility | Low CI | High CI | p-value | SD | Low CI | High CI | p-value |
| Opt-out | -3.94 | -4.93 | -2.95 | 0.000 | | | | |
| Vaccinate at pharmacy vs. health center | 0.12 | -0.38 | 0.62 | 0.637 | -0.53 | -1.42 | 0.36 | 0.241 |
| Vaccinate at community venue vs. health center | -0.49 | -0.93 | -0.06 | 0.027 | -0.02 | -0.82 | 0.79 | 0.970 |
| Vaccinate at home vs. health center | -0.98 | -1.62 | -0.34 | 0.003 | 1.21 | 0.56 | 1.85 | 0.000 |
| Vaccinate at mass site vs. health center | -0.12 | -0.73 | 0.49 | 0.701 | 0.73 | -0.21 | 1.68 | 0.129 |
| Wait for 1 hr vs. immediate service | -1.54 | -2.16 | -0.93 | 0.000 | 0.15 | -0.31 | 0.62 | 0.517 |
| Wait for 2 hrs vs. immediate service | -2.42 | -3.10 | -1.75 | 0.000 | -0.70 | -1.43 | 0.03 | 0.060 |
| Phone vs. online appointment booking | 0.48 | 0.01 | 0.95 | 0.045 | 0.77 | -0.17 | 1.71 | 0.110 |
| Drop in (no booking) vs. online appointment booking | -0.19 | -0.56 | 0.19 | 0.326 | 0.53 | -0.07 | 1.13 | 0.083 |
| Vaccinate annually vs. once | 0.14 | -0.26 | 0.54 | 0.505 | 1.01 | 0.43 | 1.59 | 0.001 |
| Enforcement for air travel vs. no enforcement | 0.44 | 0.03 | 0.84 | 0.035 | -0.19 | -1.52 | 1.13 | 0.776 |
| Enforcement for work/school vs. no enforcement | 0.68 | 0.07 | 1.30 | 0.030 | -1.29 | -2.24 | -0.34 | 0.008 |
| Enforcement for recreation vs. no enforcement | 1.24 | 0.60 | 1.89 | 0.000 | 1.24 | 0.36 | 2.11 | 0.006 |
| A few in the community vaccinated vs. no one | 0.65 | 0.23 | 1.08 | 0.003 | -0.46 | -1.05 | 0.13 | 0.126 |
| Almost everyone in the community vaccinated vs. no one | 0.50 | 0.15 | 0.85 | 0.006 | 0.29 | -1.98 | 2.56 | 0.802 |
| Two vaccine doses vs. a single dose | -4.21 | -4.90 | -3.53 | 0.000 | -0.37 | -0.95 | 0.21 | 0.210 |

Table S3(b): Weighted mean preferences – “two dose” latent class group

| Attribute | Relative utilities | | | | Standard deviation | | | |
|---|---|---|---|---|---|---|---|---|
| | Utility | Low CI | High CI | p-value | SD | Low CI | High CI | p-value |
| Opt-out | -0.45 | -1.03 | 0.13 | 0.127 | | | | |
| Vaccinate at pharmacy vs. health center | -0.38 | -0.66 | -0.11 | 0.006 | -0.42 | -0.80 | -0.03 | 0.034 |
| Vaccinate at community venue vs. health center | -0.18 | -0.54 | 0.18 | 0.322 | 0.83 | 0.39 | 1.26 | 0.000 |
| Vaccinate at home vs. health center | -1.04 | -1.53 | -0.54 | 0.000 | -0.70 | -1.13 | -0.27 | 0.001 |
| Vaccinate at mass site vs. health center | -0.67 | -1.01 | -0.32 | 0.000 | -0.44 | -1.04 | 0.15 | 0.143 |
| Wait for 1 hr vs. immediate service | -0.27 | -0.60 | 0.07 | 0.117 | 0.35 | -0.15 | 0.85 | 0.173 |
| Wait for 2 hrs vs. immediate service | -0.58 | -0.92 | -0.24 | 0.001 | 0.58 | -0.02 | 1.18 | 0.058 |
| Phone vs. online appointment booking | 0.44 | 0.20 | 0.69 | 0.000 | 0.83 | 0.41 | 1.24 | 0.000 |
| Drop in (no booking) vs. online appointment booking | -0.12 | -0.38 | 0.14 | 0.365 | 0.55 | 0.21 | 0.89 | 0.002 |
| Vaccinate annually vs. once | -0.04 | -0.25 | 0.16 | 0.682 | 0.45 | 0.07 | 0.82 | 0.020 |
| Enforcement for air travel vs. no enforcement | 0.46 | 0.19 | 0.74 | 0.001 | -0.24 | -1.05 | 0.57 | 0.564 |
| Enforcement for work/school vs. no enforcement | 0.89 | 0.64 | 1.14 | 0.000 | 0.68 | 0.24 | 1.11 | 0.002 |
| Enforcement for recreation vs. no enforcement | 0.48 | 0.17 | 0.79 | 0.003 | 0.71 | 0.24 | 1.18 | 0.003 |
| A few in the community vaccinated vs. no one | 0.40 | 0.18 | 0.61 | 0.000 | 0.28 | -0.13 | 0.70 | 0.178 |
| Almost everyone in the community vaccinated vs. no one | 0.13 | -0.07 | 0.33 | 0.198 | 0.01 | -0.42 | 0.44 | 0.972 |
| Two vaccine doses vs. a single dose | 1.72 | 1.49 | 1.95 | 0.000 | 0.72 | 0.42 | 1.02 | 0.000 |

Table S2(b): Weighted mean preferences – "vaccinate once" latent class group

| Attribute | Relative utilities | | | | Standard deviation | | | |
|---|---|---|---|---|---|---|---|---|
| | Utility | Low CI | High CI | p-value | SD | Low CI | High CI | p-value |
| Opt-out | -3.34 | -3.84 | -2.83 | 0.000 | | | | |
| Vaccinate at pharmacy vs. health center | 0.05 | -0.27 | 0.37 | 0.773 | 0.47 | -0.44 | 1.38 | 0.310 |
| Vaccinate at community venue vs. health center | -0.15 | -0.48 | 0.17 | 0.357 | 0.71 | 0.10 | 1.32 | 0.023 |
| Vaccinate at home vs. health center | -0.37 | -0.85 | 0.12 | 0.136 | 1.33 | 0.88 | 1.78 | 0.000 |
| Vaccinate at mass site vs. health center | -0.47 | -0.81 | -0.14 | 0.005 | 0.96 | 0.51 | 1.41 | 0.000 |
| Wait for 1 hr vs. immediate service | -0.72 | -1.05 | -0.38 | 0.000 | 0.53 | 0.11 | 0.95 | 0.014 |
| Wait for 2 hrs vs. immediate service | -1.02 | -1.31 | -0.72 | 0.000 | 0.93 | 0.54 | 1.33 | 0.000 |
| Phone vs. online appointment booking | 0.27 | 0.02 | 0.53 | 0.037 | 0.96 | 0.55 | 1.36 | 0.000 |
| Drop in (no booking) vs. online appointment booking | -0.02 | -0.28 | 0.25 | 0.908 | 1.02 | 0.59 | 1.45 | 0.000 |
| Vaccinate annually vs. once | -3.88 | -4.18 | -3.57 | 0.000 | 0.18 | -0.12 | 0.48 | 0.245 |
| Enforcement for air travel vs. no enforcement | -0.15 | -0.41 | 0.12 | 0.270 | 0.66 | 0.25 | 1.07 | 0.002 |
| Enforcement for work/school vs. no enforcement | -0.50 | -0.79 | -0.21 | 0.001 | 0.53 | 0.18 | 0.87 | 0.003 |
| Enforcement for recreation vs. no enforcement | -0.34 | -0.63 | -0.05 | 0.022 | 0.77 | 0.23 | 1.31 | 0.005 |
| A few in the community vaccinated vs. no one | 0.50 | 0.32 | 0.68 | 0.000 | 0.38 | -0.17 | 0.93 | 0.172 |
| Almost everyone in the community vaccinated vs. no one | 0.69 | 0.49 | 0.89 | 0.000 | 0.83 | 0.46 | 1.19 | 0.000 |
| Two vaccine doses vs. a single dose | -0.70 | -0.89 | -0.51 | 0.000 | 0.68 | 0.05 | 1.32 | 0.034 |

Table S2(c): Weighted mean preferences – “immediate service” latent class group

| Attribute | Relative utilities | | | | Standard deviation | | | |
|---|---|---|---|---|---|---|---|---|
| | Utility | Low CI | High CI | p-value | SD | Low CI | High CI | p-value |
| Opt-out | -3.87 | -4.71 | -3.02 | 0.000 | | | | |
| Vaccinate at pharmacy vs. health center | 0.02 | -0.50 | 0.54 | 0.937 | -1.07 | -2.05 | -0.09 | 0.032 |
| Vaccinate at community venue vs. health center | 0.00 | -0.39 | 0.39 | 0.999 | 1.23 | 0.55 | 1.91 | 0.000 |
| Vaccinate at home vs. health center | 0.72 | -0.10 | 1.53 | 0.085 | 1.25 | 0.36 | 2.13 | 0.006 |
| Vaccinate at mass site vs. health center | -1.87 | -2.55 | -1.19 | 0.000 | -0.62 | -1.79 | 0.56 | 0.304 |
| Wait for 1 hr vs. immediate service | -2.21 | -2.75 | -1.67 | 0.000 | 0.95 | 0.58 | 1.32 | 0.000 |
| Wait for 2 hrs vs. immediate service | -4.51 | -5.21 | -3.80 | 0.000 | 0.30 | -0.12 | 0.73 | 0.164 |
| Phone vs. online appointment booking | -0.18 | -0.53 | 0.18 | 0.328 | 0.68 | 0.28 | 1.08 | 0.001 |
| Drop in (no booking) vs. online appointment booking | 0.45 | 0.08 | 0.81 | 0.017 | -0.75 | -1.19 | -0.31 | 0.001 |
| Vaccinate annually vs. once | -0.16 | -0.52 | 0.19 | 0.369 | 0.87 | 0.40 | 1.34 | 0.000 |
| Enforcement for air travel vs. no enforcement | -0.49 | -0.94 | -0.04 | 0.034 | 0.67 | 0.00 | 1.34 | 0.051 |
| Enforcement for work/school vs. no enforcement | -0.59 | -1.02 | -0.16 | 0.007 | 0.76 | 0.29 | 1.23 | 0.002 |
| Enforcement for recreation vs. no enforcement | 0.10 | -0.29 | 0.49 | 0.619 | 0.56 | 0.12 | 1.01 | 0.013 |
| A few in the community vaccinated vs. no one | 0.60 | 0.27 | 0.93 | 0.000 | 0.78 | 0.23 | 1.33 | 0.005 |
| Almost everyone in the community vaccinated vs. no one | 0.70 | 0.35 | 1.05 | 0.000 | 1.06 | 0.61 | 1.51 | 0.000 |
| Two vaccine doses vs. a single dose | -0.27 | -0.55 | 0.01 | 0.063 | 0.89 | 0.57 | 1.21 | 0.000 |

Table S2(d): Weighted mean preferences – “social proof” latent class group

| Attribute | Relative utilities | | | | Standard deviation | | | |
|---|---|---|---|---|---|---|---|---|
| | Utility | Low CI | High CI | p-value | SD | Low CI | High CI | p-value |
| Opt-out | 0.77 | 0.24 | 1.30 | | 0.86 | 0.12 | 1.60 | |
| Vaccinate at pharmacy vs. health center | -0.11 | -0.45 | 0.22 | | 1.01 | 0.47 | 1.56 | |
| Vaccinate at community venue vs. health center | 0.03 | -0.36 | 0.42 | | 0.49 | -1.19 | 2.17 | |
| Vaccinate at home vs. health center | -0.39 | -0.89 | 0.11 | | -0.51 | -1.26 | 0.23 | |
| Vaccinate at mass site vs. health center | -0.74 | -1.10 | -0.38 | | 0.94 | 0.47 | 1.42 | |
| Wait for 1 hr vs. immediate service | 0.17 | -0.18 | 0.53 | | -0.61 | -1.09 | -0.14 | |
| Wait for 2 hrs vs. immediate service | -0.11 | -0.42 | 0.21 | | 0.86 | 0.49 | 1.22 | |
| Phone vs. online appointment booking | 0.10 | -0.13 | 0.33 | | 0.81 | 0.41 | 1.22 | |
| Drop in (no booking) vs. online appointment booking | 0.55 | 0.28 | 0.81 | | 0.48 | 0.01 | 0.95 | |
| Vaccinate annually vs. once | -0.41 | -0.58 | -0.25 | | 0.69 | 0.18 | 1.21 | |
| Enforcement for air travel vs. no enforcement | 0.65 | 0.37 | 0.93 | | 0.36 | -0.50 | 1.21 | |
| Enforcement for work/school vs. no enforcement | 0.61 | 0.31 | 0.90 | | 0.34 | -0.39 | 1.07 | |
| Enforcement for recreation vs. no enforcement | 0.85 | 0.56 | 1.14 | | 0.78 | 0.40 | 1.17 | |
| A few in the community vaccinated vs. no one | 2.64 | 2.26 | 3.02 | | -0.75 | -1.16 | -0.33 | |
| Almost everyone in the community vaccinated vs. no one | 3.73 | 3.31 | 4.15 | | -0.10 | -1.08 | 0.88 | |
| Two vaccine doses vs. a single dose | -0.47 | -0.66 | -0.27 | | 0.86 | 0.12 | 1.60 | |

Table S2(e): Weighted mean preferences – “control averse” latent class group

| Attribute | Relative utilities | | | | Standard deviation | | | |
|---|---|---|---|---|---|---|---|---|
| | Utility | Low CI | High CI | p-value | SD | Low CI | High CI | p-value |
| Opt-out | -1.81 | -2.14 | -1.47 | 0.000 | | | | |
| Vaccinate at pharmacy vs. health center | 0.34 | 0.15 | 0.53 | 0.000 | 0.87 | 0.50 | 1.23 | 0.000 |
| Vaccinate at community venue vs. health center | -0.06 | -0.22 | 0.09 | 0.431 | 0.51 | -0.08 | 1.10 | 0.092 |
| Vaccinate at home vs. health center | 0.24 | -0.04 | 0.52 | 0.088 | -0.73 | -1.03 | -0.43 | 0.000 |
| Vaccinate at mass site vs. health center | 0.04 | -0.13 | 0.20 | 0.657 | 0.68 | 0.28 | 1.08 | 0.001 |
| Wait for 1 hr vs. immediate service | 0.15 | -0.02 | 0.32 | 0.091 | 0.67 | 0.46 | 0.89 | 0.000 |
| Wait for 2 hrs vs. immediate service | -0.03 | -0.20 | 0.13 | 0.683 | 0.62 | 0.28 | 0.96 | 0.000 |
| Phone vs. online appointment booking | -0.49 | -0.64 | -0.34 | 0.000 | 0.26 | -0.89 | 1.40 | 0.659 |
| Drop in (no booking) vs. online appointment booking | -0.10 | -0.24 | 0.05 | 0.201 | 0.47 | 0.09 | 0.86 | 0.016 |
| Vaccinate annually vs. once | -0.15 | -0.26 | -0.04 | 0.010 | 0.40 | 0.20 | 0.59 | 0.000 |
| Enforcement for air travel vs. no enforcement | -0.69 | -0.86 | -0.52 | 0.000 | 0.82 | 0.61 | 1.03 | 0.000 |
| Enforcement for work/school vs. no enforcement | -0.86 | -1.03 | -0.69 | 0.000 | -0.47 | -0.94 | 0.01 | 0.053 |
| Enforcement for recreation vs. no enforcement | -0.55 | -0.71 | -0.39 | 0.000 | 0.62 | 0.38 | 0.86 | 0.000 |
| A few in the community vaccinated vs. no one | -0.13 | -0.26 | -0.01 | 0.035 | 0.29 | -0.17 | 0.75 | 0.221 |
| Almost everyone in the community vaccinated vs. no one | -0.07 | -0.20 | 0.06 | 0.297 | 0.19 | -0.48 | 0.86 | 0.582 |
| Two vaccine doses vs. a single dose | -0.33 | -0.43 | -0.24 | 0.000 | 0.46 | 0.19 | 0.74 | 0.001 |

Table S4: Marginal probability of latent class group membership

| | Control averse | | | Immediate service | | | Vaccine features | | | Social proof | | |
|---|---|---|---|---|---|---|---|---|---|---|---|---|
| Characteristic | Marginal Probability | Low CI | High CI | Marginal Probability | Low CI | High CI | Marginal Probability | Low CI | High CI | Marginal Probability | Low CI | High CI |
| Age 18-24 yrs | 35.2% | 26.2% | 44.3% | 9.5% | 3.1% | 15.8% | 37.7% | 27.3% | 48.0% | 17.6% | 10.5% | 24.7% |
| Age 25-34 yrs | 42.7% | 34.1% | 51.3% | 4.6% | 1.5% | 7.6% | 39.5% | 31.6% | 47.4% | 13.3% | 8.7% | 17.9% |
| Age 35-44 yrs | 41.4% | 35.3% | 47.4% | 8.9% | 4.5% | 13.4% | 37.7% | 31.6% | 43.9% | 12.0% | 8.3% | 15.7% |
| Age 45-54 yrs | 30.9% | 24.7% | 37.2% | 5.9% | 3.5% | 8.3% | 49.0% | 41.8% | 56.3% | 14.1% | 9.6% | 18.7% |
| Age 55-64 yrs | 29.3% | 24.5% | 34.0% | 9.7% | 7.2% | 12.3% | 48.6% | 43.4% | 53.9% | 12.3% | 9.0% | 15.7% |
| Age 65+ yrs | 20.1% | 15.8% | 24.4% | 9.9% | 6.4% | 13.4% | 60.4% | 54.8% | 66.0% | 9.6% | 6.4% | 12.8% |
| Black/ African American | 38.4% | 32.9% | 44.0% | 6.0% | 3.6% | 8.3% | 47.7% | 41.8% | 53.5% | 7.9% | 5.4% | 10.5% |
| White | 31.5% | 28.3% | 34.6% | 8.5% | 6.6% | 10.3% | 46.9% | 43.6% | 50.2% | 13.1% | 11.1% | 15.2% |
| Asian | 25.5% | 17.7% | 33.3% | 9.7% | 4.3% | 15.0% | 49.3% | 39.8% | 58.8% | 15.5% | 9.3% | 21.7% |
| Other race | 43.5% | 31.3% | 55.7% | 5.4% | 0.0% | 11.5% | 37.8% | 25.5% | 50.1% | 13.3% | 3.3% | 23.3% |
| Democrat | 31.8% | 27.9% | 35.6% | 8.3% | 6.1% | 10.5% | 44.7% | 40.8% | 48.6% | 15.2% | 12.2% | 18.2% |
| Republican | 33.7% | 28.4% | 39.0% | 7.1% | 5.0% | 9.1% | 51.7% | 45.8% | 57.7% | 7.5% | 5.2% | 9.8% |
| Other political group | 32.7% | 27.7% | 37.7% | 8.9% | 5.3% | 12.6% | 45.5% | 40.0% | 51.0% | 12.9% | 9.6% | 16.1% |
| Already/definitely vaccinate | 29.1% | 26.0% | 32.2% | 9.0% | 7.2% | 10.9% | 48.6% | 45.2% | 52.0% | 13.3% | 11.2% | 15.3% |
| Probably vaccinate | 39.0% | 31.7% | 46.4% | 5.3% | 2.7% | 8.0% | 45.1% | 37.5% | 52.8% | 10.5% | 6.6% | 14.4% |
| Probably not vaccinate | 40.0% | 31.2% | 48.9% | 6.0% | 2.7% | 9.4% | 38.9% | 30.0% | 47.7% | 15.1% | 8.0% | 22.1% |
| Definitely not vaccinate | 52.1% | 40.0% | 64.3% | 4.6% | 0.4% | 8.7% | 38.7% | 26.6% | 50.9% | 4.6% | 0.4% | 8.7% |

| Table S5: Pulse Survey population percentages as applied to reweight DCE study sample | | | | | |
|---|---|---|---|---|---|
| Race | Age Group | Gender | Education | Vaccine Status | Percentage in US population |
| White | 18-24 | Male | High school or less | Definitely get a vaccine | 0.88059670 |
| White | 18-24 | Male | High school or less | Probably get a vaccine | 0.41185856 |
| White | 18-24 | Male | High school or less | Probably NOT get a vaccine | 0.20597842 |
| White | 18-24 | Male | High school or less | Definitely NOT get a vaccine | 0.20889288 |
| White | 18-24 | Male | High school or less | Already vaccinated | 0.14291063 |
| White | 18-24 | Male | Incomplete college/Associate degree | Definitely get a vaccine | 0.65890247 |
| White | 18-24 | Male | Incomplete college/Associate degree | Probably get a vaccine | 0.27298173 |
| White | 18-24 | Male | Incomplete college/Associate degree | Probably NOT get a vaccine | 0.11789024 |
| White | 18-24 | Male | Incomplete college/Associate degree | Definitely NOT get a vaccine | 0.08753720 |
| White | 18-24 | Male | Incomplete college/Associate degree | Already vaccinated | 0.21155982 |
| White | 18-24 | Male | Bachelor degree or higher | Definitely get a vaccine | 0.18038267 |
| White | 18-24 | Male | Bachelor degree or higher | Probably get a vaccine | 0.02283200 |
| White | 18-24 | Male | Bachelor degree or higher | Probably NOT get a vaccine | 0.00799903 |
| White | 18-24 | Male | Bachelor degree or higher | Definitely NOT get a vaccine | 0.01547569 |
| White | 18-24 | Male | Bachelor degree or higher | Already vaccinated | 0.07429212 |
| White | 18-24 | Female | High school or less | Definitely get a vaccine | 0.41138631 |
| White | 18-24 | Female | High school or less | Probably get a vaccine | 0.26984257 |
| White | 18-24 | Female | High school or less | Probably NOT get a vaccine | 0.12144046 |
| White | 18-24 | Female | High school or less | Definitely NOT get a vaccine | 0.10323959 |
| White | 18-24 | Female | High school or less | Already vaccinated | 0.12998548 |
| White | 18-24 | Female | Incomplete college/Associate degree | Definitely get a vaccine | 0.69271505 |
| White | 18-24 | Female | Incomplete college/Associate degree | Probably get a vaccine | 0.17073895 |
| White | 18-24 | Female | Incomplete college/Associate degree | Probably NOT get a vaccine | 0.17620140 |
| White | 18-24 | Female | Incomplete college/Associate degree | Definitely NOT get a vaccine | 0.09844478 |
| White | 18-24 | Female | Incomplete college/Associate degree | Already vaccinated | 0.43343481 |
| White | 18-24 | Female | Bachelor degree or higher | Definitely get a vaccine | 0.12168144 |
| White | 18-24 | Female | Bachelor degree or higher | Probably get a vaccine | 0.03296001 |
| White | 18-24 | Female | Bachelor degree or higher | Probably NOT get a vaccine | 0.03025279 |
| White | 18-24 | Female | Bachelor degree or higher | Definitely NOT get a vaccine | 0.00429185 |
| White | 18-24 | Female | Bachelor degree or higher | Already vaccinated | 0.18734966 |
| White | 25-34 | Male | High school or less | Definitely get a vaccine | 0.55930632 |
| White | 25-34 | Male | High school or less | Probably get a vaccine | 0.43332657 |
| White | 25-34 | Male | High school or less | Probably NOT get a vaccine | 0.37375864 |
| White | 25-34 | Male | High school or less | Definitely NOT get a vaccine | 0.45896083 |
| White | 25-34 | Male | High school or less | Already vaccinated | 0.21719511 |
| White | 25-34 | Male | Incomplete college/Associate degree | Definitely get a vaccine | 0.77293873 |
| White | 25-34 | Male | Incomplete college/Associate degree | Probably get a vaccine | 0.33312801 |
| White | 25-34 | Male | Incomplete college/Associate degree | Probably NOT get a vaccine | 0.18209177 |
| White | 25-34 | Male | Incomplete college/Associate degree | Definitely NOT get a vaccine | 0.23028831 |
| White | 25-34 | Male | Incomplete college/Associate degree | Already vaccinated | 0.58378553 |
| White | 25-34 | Male | Bachelor degree or higher | Definitely get a vaccine | 0.98175013 |
| White | 25-34 | Male | Bachelor degree or higher | Probably get a vaccine | 0.17452127 |
| White | 25-34 | Male | Bachelor degree or higher | Probably NOT get a vaccine | 0.11903349 |
| White | 25-34 | Male | Bachelor degree or higher | Definitely NOT get a vaccine | 0.05970662 |

| | | | | | |
|---|---|---|---|---|---|
| White | 25-34 | Male | Bachelor degree or higher | Already vaccinated | 0.84533185 |
| White | 25-34 | Female | High school or less | Definitely get a vaccine | 0.44100893 |
| White | 25-34 | Female | High school or less | Probably get a vaccine | 0.36426613 |
| White | 25-34 | Female | High school or less | Probably NOT get a vaccine | 0.25183254 |
| White | 25-34 | Female | High school or less | Definitely NOT get a vaccine | 0.31789559 |
| White | 25-34 | Female | High school or less | Already vaccinated | 0.34762943 |
| White | 25-34 | Female | Incomplete college/Associate degree | Definitely get a vaccine | 0.63471496 |
| White | 25-34 | Female | Incomplete college/Associate degree | Probably get a vaccine | 0.43889531 |
| White | 25-34 | Female | Incomplete college/Associate degree | Probably NOT get a vaccine | 0.30291691 |
| White | 25-34 | Female | Incomplete college/Associate degree | Definitely NOT get a vaccine | 0.29751211 |
| White | 25-34 | Female | Incomplete college/Associate degree | Already vaccinated | 0.66430008 |
| White | 25-34 | Female | Bachelor degree or higher | Definitely get a vaccine | 0.83233625 |
| White | 25-34 | Female | Bachelor degree or higher | Probably get a vaccine | 0.22707644 |
| White | 25-34 | Female | Bachelor degree or higher | Probably NOT get a vaccine | 0.20688841 |
| White | 25-34 | Female | Bachelor degree or higher | Definitely NOT get a vaccine | 0.09568925 |
| White | 25-34 | Female | Bachelor degree or higher | Already vaccinated | 1.50736952 |
| White | 35-44 | Male | High school or less | Definitely get a vaccine | 0.80106801 |
| White | 35-44 | Male | High school or less | Probably get a vaccine | 0.39562032 |
| White | 35-44 | Male | High school or less | Probably NOT get a vaccine | 0.27381086 |
| White | 35-44 | Male | High school or less | Definitely NOT get a vaccine | 0.37029043 |
| White | 35-44 | Male | High school or less | Already vaccinated | 0.55618668 |
| White | 35-44 | Male | Incomplete college/Associate degree | Definitely get a vaccine | 0.64574122 |
| White | 35-44 | Male | Incomplete college/Associate degree | Probably get a vaccine | 0.25875312 |
| White | 35-44 | Male | Incomplete college/Associate degree | Probably NOT get a vaccine | 0.21732798 |
| White | 35-44 | Male | Incomplete college/Associate degree | Definitely NOT get a vaccine | 0.24663784 |
| White | 35-44 | Male | Incomplete college/Associate degree | Already vaccinated | 0.64785451 |
| White | 35-44 | Male | Bachelor degree or higher | Definitely get a vaccine | 0.72330636 |
| White | 35-44 | Male | Bachelor degree or higher | Probably get a vaccine | 0.17681870 |
| White | 35-44 | Male | Bachelor degree or higher | Probably NOT get a vaccine | 0.11164686 |
| White | 35-44 | Male | Bachelor degree or higher | Definitely NOT get a vaccine | 0.12200039 |
| White | 35-44 | Male | Bachelor degree or higher | Already vaccinated | 1.06056833 |
| White | 35-44 | Female | High school or less | Definitely get a vaccine | 0.47539768 |
| White | 35-44 | Female | High school or less | Probably get a vaccine | 0.46099666 |
| White | 35-44 | Female | High school or less | Probably NOT get a vaccine | 0.24208607 |
| White | 35-44 | Female | High school or less | Definitely NOT get a vaccine | 0.30721790 |
| White | 35-44 | Female | High school or less | Already vaccinated | 0.43671286 |
| White | 35-44 | Female | Incomplete college/Associate degree | Definitely get a vaccine | 0.38280156 |
| White | 35-44 | Female | Incomplete college/Associate degree | Probably get a vaccine | 0.29467374 |
| White | 35-44 | Female | Incomplete college/Associate degree | Probably NOT get a vaccine | 0.25390655 |
| White | 35-44 | Female | Incomplete college/Associate degree | Definitely NOT get a vaccine | 0.24215461 |
| White | 35-44 | Female | Incomplete college/Associate degree | Already vaccinated | 0.63146037 |
| White | 35-44 | Female | Bachelor degree or higher | Definitely get a vaccine | 0.65702331 |
| White | 35-44 | Female | Bachelor degree or higher | Probably get a vaccine | 0.18878050 |
| White | 35-44 | Female | Bachelor degree or higher | Probably NOT get a vaccine | 0.16097838 |
| White | 35-44 | Female | Bachelor degree or higher | Definitely NOT get a vaccine | 0.10673342 |
| White | 35-44 | Female | Bachelor degree or higher | Already vaccinated | 1.43354511 |

| | | | | | |
|---|---|---|---|---|---|
| White | 45-54 | Male | High school or less | Definitely get a vaccine | 0.78012407 |
| White | 45-54 | Male | High school or less | Probably get a vaccine | 0.33711344 |
| White | 45-54 | Male | High school or less | Probably NOT get a vaccine | 0.21099958 |
| White | 45-54 | Male | High school or less | Definitely NOT get a vaccine | 0.38228554 |
| White | 45-54 | Male | High school or less | Already vaccinated | 0.73247677 |
| White | 45-54 | Male | Incomplete college/Associate degree | Definitely get a vaccine | 0.42353806 |
| White | 45-54 | Male | Incomplete college/Associate degree | Probably get a vaccine | 0.23593360 |
| White | 45-54 | Male | Incomplete college/Associate degree | Probably NOT get a vaccine | 0.14247265 |
| White | 45-54 | Male | Incomplete college/Associate degree | Definitely NOT get a vaccine | 0.14490201 |
| White | 45-54 | Male | Incomplete college/Associate degree | Already vaccinated | 0.66418236 |
| White | 45-54 | Male | Bachelor degree or higher | Definitely get a vaccine | 0.60467201 |
| White | 45-54 | Male | Bachelor degree or higher | Probably get a vaccine | 0.19385101 |
| White | 45-54 | Male | Bachelor degree or higher | Probably NOT get a vaccine | 0.06460436 |
| White | 45-54 | Male | Bachelor degree or higher | Definitely NOT get a vaccine | 0.07184025 |
| White | 45-54 | Male | Bachelor degree or higher | Already vaccinated | 0.97267097 |
| White | 45-54 | Female | High school or less | Definitely get a vaccine | 0.71404314 |
| White | 45-54 | Female | High school or less | Probably get a vaccine | 0.39102462 |
| White | 45-54 | Female | High school or less | Probably NOT get a vaccine | 0.24584676 |
| White | 45-54 | Female | High school or less | Definitely NOT get a vaccine | 0.24102242 |
| White | 45-54 | Female | High school or less | Already vaccinated | 0.58880097 |
| White | 45-54 | Female | Incomplete college/Associate degree | Definitely get a vaccine | 0.38530064 |
| White | 45-54 | Female | Incomplete college/Associate degree | Probably get a vaccine | 0.22766306 |
| White | 45-54 | Female | Incomplete college/Associate degree | Probably NOT get a vaccine | 0.18935494 |
| White | 45-54 | Female | Incomplete college/Associate degree | Definitely NOT get a vaccine | 0.18744701 |
| White | 45-54 | Female | Incomplete college/Associate degree | Already vaccinated | 0.81366622 |
| White | 45-54 | Female | Bachelor degree or higher | Definitely get a vaccine | 0.48854232 |
| White | 45-54 | Female | Bachelor degree or higher | Probably get a vaccine | 0.16724294 |
| White | 45-54 | Female | Bachelor degree or higher | Probably NOT get a vaccine | 0.11898842 |
| White | 45-54 | Female | Bachelor degree or higher | Definitely NOT get a vaccine | 0.08746195 |
| White | 45-54 | Female | Bachelor degree or higher | Already vaccinated | 1.23043501 |
| White | 55-64 | Male | High school or less | Definitely get a vaccine | 0.73256952 |
| White | 55-64 | Male | High school or less | Probably get a vaccine | 0.40250343 |
| White | 55-64 | Male | High school or less | Probably NOT get a vaccine | 0.23209365 |
| White | 55-64 | Male | High school or less | Definitely NOT get a vaccine | 0.21451372 |
| White | 55-64 | Male | High school or less | Already vaccinated | 1.28672719 |
| White | 55-64 | Male | Incomplete college/Associate degree | Definitely get a vaccine | 0.43482670 |
| White | 55-64 | Male | Incomplete college/Associate degree | Probably get a vaccine | 0.19126257 |
| White | 55-64 | Male | Incomplete college/Associate degree | Probably NOT get a vaccine | 0.11919882 |
| White | 55-64 | Male | Incomplete college/Associate degree | Definitely NOT get a vaccine | 0.15724355 |
| White | 55-64 | Male | Incomplete college/Associate degree | Already vaccinated | 0.94029742 |
| White | 55-64 | Male | Bachelor degree or higher | Definitely get a vaccine | 0.48590744 |
| White | 55-64 | Male | Bachelor degree or higher | Probably get a vaccine | 0.11547807 |
| White | 55-64 | Male | Bachelor degree or higher | Probably NOT get a vaccine | 0.05247556 |
| White | 55-64 | Male | Bachelor degree or higher | Definitely NOT get a vaccine | 0.06422006 |
| White | 55-64 | Male | Bachelor degree or higher | Already vaccinated | 1.12754321 |
| White | 55-64 | Female | High school or less | Definitely get a vaccine | 0.66534728 |

| | | | | | |
|---|---|---|---|---|---|
| White | 55-64 | Female | High school or less | Probably get a vaccine | 0.36104518 |
| White | 55-64 | Female | High school or less | Probably NOT get a vaccine | 0.22285943 |
| White | 55-64 | Female | High school or less | Definitely NOT get a vaccine | 0.16690432 |
| White | 55-64 | Female | High school or less | Already vaccinated | 1.28392661 |
| White | 55-64 | Female | Incomplete college/Associate degree | Definitely get a vaccine | 0.47443166 |
| White | 55-64 | Female | Incomplete college/Associate degree | Probably get a vaccine | 0.22045954 |
| White | 55-64 | Female | Incomplete college/Associate degree | Probably NOT get a vaccine | 0.14498445 |
| White | 55-64 | Female | Incomplete college/Associate degree | Definitely NOT get a vaccine | 0.11059872 |
| White | 55-64 | Female | Incomplete college/Associate degree | Already vaccinated | 1.15686619 |
| White | 55-64 | Female | Bachelor degree or higher | Definitely get a vaccine | 0.42633542 |
| White | 55-64 | Female | Bachelor degree or higher | Probably get a vaccine | 0.10102709 |
| White | 55-64 | Female | Bachelor degree or higher | Probably NOT get a vaccine | 0.07827702 |
| White | 55-64 | Female | Bachelor degree or higher | Definitely NOT get a vaccine | 0.05280546 |
| White | 55-64 | Female | Bachelor degree or higher | Already vaccinated | 1.33369315 |
| White | 65+ | Male | High school or less | Definitely get a vaccine | 0.39842489 |
| White | 65+ | Male | High school or less | Probably get a vaccine | 0.14682660 |
| White | 65+ | Male | High school or less | Probably NOT get a vaccine | 0.17538460 |
| White | 65+ | Male | High school or less | Definitely NOT get a vaccine | 0.12438075 |
| White | 65+ | Male | High school or less | Already vaccinated | 2.46911931 |
| White | 65+ | Male | Incomplete college/Associate degree | Definitely get a vaccine | 0.17081732 |
| White | 65+ | Male | Incomplete college/Associate degree | Probably get a vaccine | 0.08464184 |
| White | 65+ | Male | Incomplete college/Associate degree | Probably NOT get a vaccine | 0.05811615 |
| White | 65+ | Male | Incomplete college/Associate degree | Definitely NOT get a vaccine | 0.07587925 |
| White | 65+ | Male | Incomplete college/Associate degree | Already vaccinated | 1.75297391 |
| White | 65+ | Male | Bachelor degree or higher | Definitely get a vaccine | 0.14756335 |
| White | 65+ | Male | Bachelor degree or higher | Probably get a vaccine | 0.05802047 |
| White | 65+ | Male | Bachelor degree or higher | Probably NOT get a vaccine | 0.04194709 |
| White | 65+ | Male | Bachelor degree or higher | Definitely NOT get a vaccine | 0.04490284 |
| White | 65+ | Male | Bachelor degree or higher | Already vaccinated | 2.45612597 |
| White | 65+ | Female | High school or less | Definitely get a vaccine | 0.41485068 |
| White | 65+ | Female | High school or less | Probably get a vaccine | 0.26718476 |
| White | 65+ | Female | High school or less | Probably NOT get a vaccine | 0.21084957 |
| White | 65+ | Female | High school or less | Definitely NOT get a vaccine | 0.16612864 |
| White | 65+ | Female | High school or less | Already vaccinated | 3.68626857 |
| White | 65+ | Female | Incomplete college/Associate degree | Definitely get a vaccine | 0.18964103 |
| White | 65+ | Female | Incomplete college/Associate degree | Probably get a vaccine | 0.10361837 |
| White | 65+ | Female | Incomplete college/Associate degree | Probably NOT get a vaccine | 0.10089190 |
| White | 65+ | Female | Incomplete college/Associate degree | Definitely NOT get a vaccine | 0.10063599 |
| White | 65+ | Female | Incomplete college/Associate degree | Already vaccinated | 2.16050768 |
| White | 65+ | Female | Bachelor degree or higher | Definitely get a vaccine | 0.12978722 |
| White | 65+ | Female | Bachelor degree or higher | Probably get a vaccine | 0.05568712 |
| White | 65+ | Female | Bachelor degree or higher | Probably NOT get a vaccine | 0.04837983 |
| White | 65+ | Female | Bachelor degree or higher | Definitely NOT get a vaccine | 0.04076084 |
| White | 65+ | Female | Bachelor degree or higher | Already vaccinated | 2.12997341 |
| Black | 18-24 | Male | High school or less | Definitely get a vaccine | 0.04254603 |
| Black | 18-24 | Male | High school or less | Probably get a vaccine | 0.06432409 |

| | | | | | |
|---|---|---|---|---|---|
| Black | 18-24 | Male | High school or less | Probably NOT get a vaccine | 0.00000000 |
| Black | 18-24 | Male | High school or less | Definitely NOT get a vaccine | 0.03150065 |
| Black | 18-24 | Male | High school or less | Already vaccinated | 0.03623263 |
| Black | 18-24 | Male | Incomplete college/Associate degree | Definitely get a vaccine | 0.04990794 |
| Black | 18-24 | Male | Incomplete college/Associate degree | Probably get a vaccine | 0.03823696 |
| Black | 18-24 | Male | Incomplete college/Associate degree | Probably NOT get a vaccine | 0.04109951 |
| Black | 18-24 | Male | Incomplete college/Associate degree | Definitely NOT get a vaccine | 0.01017068 |
| Black | 18-24 | Male | Incomplete college/Associate degree | Already vaccinated | 0.01273386 |
| Black | 18-24 | Male | Bachelor degree or higher | Definitely get a vaccine | 0.01869441 |
| Black | 18-24 | Male | Bachelor degree or higher | Probably get a vaccine | 0.00000000 |
| Black | 18-24 | Male | Bachelor degree or higher | Probably NOT get a vaccine | 0.00297286 |
| Black | 18-24 | Male | Bachelor degree or higher | Definitely NOT get a vaccine | 0.00000000 |
| Black | 18-24 | Male | Bachelor degree or higher | Already vaccinated | 0.00203632 |
| Black | 18-24 | Female | High school or less | Definitely get a vaccine | 0.04540189 |
| Black | 18-24 | Female | High school or less | Probably get a vaccine | 0.02947550 |
| Black | 18-24 | Female | High school or less | Probably NOT get a vaccine | 0.02006121 |
| Black | 18-24 | Female | High school or less | Definitely NOT get a vaccine | 0.02300096 |
| Black | 18-24 | Female | High school or less | Already vaccinated | 0.00391023 |
| Black | 18-24 | Female | Incomplete college/Associate degree | Definitely get a vaccine | 0.04535841 |
| Black | 18-24 | Female | Incomplete college/Associate degree | Probably get a vaccine | 0.08136075 |
| Black | 18-24 | Female | Incomplete college/Associate degree | Probably NOT get a vaccine | 0.01870689 |
| Black | 18-24 | Female | Incomplete college/Associate degree | Definitely NOT get a vaccine | 0.02017083 |
| Black | 18-24 | Female | Incomplete college/Associate degree | Already vaccinated | 0.05532825 |
| Black | 18-24 | Female | Bachelor degree or higher | Definitely get a vaccine | 0.01165846 |
| Black | 18-24 | Female | Bachelor degree or higher | Probably get a vaccine | 0.01256022 |
| Black | 18-24 | Female | Bachelor degree or higher | Probably NOT get a vaccine | 0.01190885 |
| Black | 18-24 | Female | Bachelor degree or higher | Definitely NOT get a vaccine | 0.00000000 |
| Black | 18-24 | Female | Bachelor degree or higher | Already vaccinated | 0.00819009 |
| Black | 25-34 | Male | High school or less | Definitely get a vaccine | 0.07604944 |
| Black | 25-34 | Male | High school or less | Probably get a vaccine | 0.08116805 |
| Black | 25-34 | Male | High school or less | Probably NOT get a vaccine | 0.04534615 |
| Black | 25-34 | Male | High school or less | Definitely NOT get a vaccine | 0.10170152 |
| Black | 25-34 | Male | High school or less | Already vaccinated | 0.01981156 |
| Black | 25-34 | Male | Incomplete college/Associate degree | Definitely get a vaccine | 0.04979984 |
| Black | 25-34 | Male | Incomplete college/Associate degree | Probably get a vaccine | 0.11900820 |
| Black | 25-34 | Male | Incomplete college/Associate degree | Probably NOT get a vaccine | 0.03680303 |
| Black | 25-34 | Male | Incomplete college/Associate degree | Definitely NOT get a vaccine | 0.00334251 |
| Black | 25-34 | Male | Incomplete college/Associate degree | Already vaccinated | 0.01193117 |
| Black | 25-34 | Male | Bachelor degree or higher | Definitely get a vaccine | 0.06161587 |
| Black | 25-34 | Male | Bachelor degree or higher | Probably get a vaccine | 0.04612500 |
| Black | 25-34 | Male | Bachelor degree or higher | Probably NOT get a vaccine | 0.00854158 |
| Black | 25-34 | Male | Bachelor degree or higher | Definitely NOT get a vaccine | 0.00036173 |
| Black | 25-34 | Male | Bachelor degree or higher | Already vaccinated | 0.06519294 |
| Black | 25-34 | Female | High school or less | Definitely get a vaccine | 0.08240709 |
| Black | 25-34 | Female | High school or less | Probably get a vaccine | 0.11121935 |
| Black | 25-34 | Female | High school or less | Probably NOT get a vaccine | 0.10904006 |

| | | | | | |
|---|---|---|---|---|---|
| Black | 25-34 | Female | High school or less | Definitely NOT get a vaccine | 0.13209657 |
| Black | 25-34 | Female | High school or less | Already vaccinated | 0.04426508 |
| Black | 25-34 | Female | Incomplete college/Associate degree | Definitely get a vaccine | 0.08719063 |
| Black | 25-34 | Female | Incomplete college/Associate degree | Probably get a vaccine | 0.09444108 |
| Black | 25-34 | Female | Incomplete college/Associate degree | Probably NOT get a vaccine | 0.10006076 |
| Black | 25-34 | Female | Incomplete college/Associate degree | Definitely NOT get a vaccine | 0.05717136 |
| Black | 25-34 | Female | Incomplete college/Associate degree | Already vaccinated | 0.09763716 |
| Black | 25-34 | Female | Bachelor degree or higher | Definitely get a vaccine | 0.07728636 |
| Black | 25-34 | Female | Bachelor degree or higher | Probably get a vaccine | 0.07221153 |
| Black | 25-34 | Female | Bachelor degree or higher | Probably NOT get a vaccine | 0.03335232 |
| Black | 25-34 | Female | Bachelor degree or higher | Definitely NOT get a vaccine | 0.02093162 |
| Black | 25-34 | Female | Bachelor degree or higher | Already vaccinated | 0.11235956 |
| Black | 35-44 | Male | High school or less | Definitely get a vaccine | 0.18304512 |
| Black | 35-44 | Male | High school or less | Probably get a vaccine | 0.08327250 |
| Black | 35-44 | Male | High school or less | Probably NOT get a vaccine | 0.10193397 |
| Black | 35-44 | Male | High school or less | Definitely NOT get a vaccine | 0.09855625 |
| Black | 35-44 | Male | High school or less | Already vaccinated | 0.08943479 |
| Black | 35-44 | Male | Incomplete college/Associate degree | Definitely get a vaccine | 0.17146583 |
| Black | 35-44 | Male | Incomplete college/Associate degree | Probably get a vaccine | 0.07702290 |
| Black | 35-44 | Male | Incomplete college/Associate degree | Probably NOT get a vaccine | 0.03318253 |
| Black | 35-44 | Male | Incomplete college/Associate degree | Definitely NOT get a vaccine | 0.06048685 |
| Black | 35-44 | Male | Incomplete college/Associate degree | Already vaccinated | 0.12700726 |
| Black | 35-44 | Male | Bachelor degree or higher | Definitely get a vaccine | 0.07688067 |
| Black | 35-44 | Male | Bachelor degree or higher | Probably get a vaccine | 0.03699781 |
| Black | 35-44 | Male | Bachelor degree or higher | Probably NOT get a vaccine | 0.01014439 |
| Black | 35-44 | Male | Bachelor degree or higher | Definitely NOT get a vaccine | 0.00784530 |
| Black | 35-44 | Male | Bachelor degree or higher | Already vaccinated | 0.08971959 |
| Black | 35-44 | Female | High school or less | Definitely get a vaccine | 0.13272803 |
| Black | 35-44 | Female | High school or less | Probably get a vaccine | 0.13793813 |
| Black | 35-44 | Female | High school or less | Probably NOT get a vaccine | 0.14260516 |
| Black | 35-44 | Female | High school or less | Definitely NOT get a vaccine | 0.11364584 |
| Black | 35-44 | Female | High school or less | Already vaccinated | 0.18334116 |
| Black | 35-44 | Female | Incomplete college/Associate degree | Definitely get a vaccine | 0.09592485 |
| Black | 35-44 | Female | Incomplete college/Associate degree | Probably get a vaccine | 0.08537077 |
| Black | 35-44 | Female | Incomplete college/Associate degree | Probably NOT get a vaccine | 0.11339032 |
| Black | 35-44 | Female | Incomplete college/Associate degree | Definitely NOT get a vaccine | 0.06999999 |
| Black | 35-44 | Female | Incomplete college/Associate degree | Already vaccinated | 0.12403256 |
| Black | 35-44 | Female | Bachelor degree or higher | Definitely get a vaccine | 0.06652909 |
| Black | 35-44 | Female | Bachelor degree or higher | Probably get a vaccine | 0.07837132 |
| Black | 35-44 | Female | Bachelor degree or higher | Probably NOT get a vaccine | 0.06699082 |
| Black | 35-44 | Female | Bachelor degree or higher | Definitely NOT get a vaccine | 0.01266304 |
| Black | 35-44 | Female | Bachelor degree or higher | Already vaccinated | 0.18936825 |
| Black | 45-54 | Male | High school or less | Definitely get a vaccine | 0.20094514 |
| Black | 45-54 | Male | High school or less | Probably get a vaccine | 0.06883197 |
| Black | 45-54 | Male | High school or less | Probably NOT get a vaccine | 0.07823318 |
| Black | 45-54 | Male | High school or less | Definitely NOT get a vaccine | 0.02786543 |

| | | | | | |
|---|---|---|---|---|---|
| Black | 45-54 | Male | High school or less | Already vaccinated | 0.15776481 |
| Black | 45-54 | Male | Incomplete college/Associate degree | Definitely get a vaccine | 0.10815254 |
| Black | 45-54 | Male | Incomplete college/Associate degree | Probably get a vaccine | 0.07793187 |
| Black | 45-54 | Male | Incomplete college/Associate degree | Probably NOT get a vaccine | 0.02135873 |
| Black | 45-54 | Male | Incomplete college/Associate degree | Definitely NOT get a vaccine | 0.01941395 |
| Black | 45-54 | Male | Incomplete college/Associate degree | Already vaccinated | 0.14445241 |
| Black | 45-54 | Male | Bachelor degree or higher | Definitely get a vaccine | 0.06964424 |
| Black | 45-54 | Male | Bachelor degree or higher | Probably get a vaccine | 0.04596581 |
| Black | 45-54 | Male | Bachelor degree or higher | Probably NOT get a vaccine | 0.00666862 |
| Black | 45-54 | Male | Bachelor degree or higher | Definitely NOT get a vaccine | 0.00118402 |
| Black | 45-54 | Male | Bachelor degree or higher | Already vaccinated | 0.12998950 |
| Black | 45-54 | Female | High school or less | Definitely get a vaccine | 0.14459036 |
| Black | 45-54 | Female | High school or less | Probably get a vaccine | 0.11687817 |
| Black | 45-54 | Female | High school or less | Probably NOT get a vaccine | 0.03644238 |
| Black | 45-54 | Female | High school or less | Definitely NOT get a vaccine | 0.03273686 |
| Black | 45-54 | Female | High school or less | Already vaccinated | 0.13599311 |
| Black | 45-54 | Female | Incomplete college/Associate degree | Definitely get a vaccine | 0.06573137 |
| Black | 45-54 | Female | Incomplete college/Associate degree | Probably get a vaccine | 0.08759151 |
| Black | 45-54 | Female | Incomplete college/Associate degree | Probably NOT get a vaccine | 0.04020254 |
| Black | 45-54 | Female | Incomplete college/Associate degree | Definitely NOT get a vaccine | 0.03154715 |
| Black | 45-54 | Female | Incomplete college/Associate degree | Already vaccinated | 0.17598599 |
| Black | 45-54 | Female | Bachelor degree or higher | Definitely get a vaccine | 0.04741024 |
| Black | 45-54 | Female | Bachelor degree or higher | Probably get a vaccine | 0.03366010 |
| Black | 45-54 | Female | Bachelor degree or higher | Probably NOT get a vaccine | 0.02546248 |
| Black | 45-54 | Female | Bachelor degree or higher | Definitely NOT get a vaccine | 0.00638455 |
| Black | 45-54 | Female | Bachelor degree or higher | Already vaccinated | 0.20653744 |
| Black | 55-64 | Male | High school or less | Definitely get a vaccine | 0.08532783 |
| Black | 55-64 | Male | High school or less | Probably get a vaccine | 0.11139014 |
| Black | 55-64 | Male | High school or less | Probably NOT get a vaccine | 0.05600410 |
| Black | 55-64 | Male | High school or less | Definitely NOT get a vaccine | 0.00908051 |
| Black | 55-64 | Male | High school or less | Already vaccinated | 0.22827449 |
| Black | 55-64 | Male | Incomplete college/Associate degree | Definitely get a vaccine | 0.07828084 |
| Black | 55-64 | Male | Incomplete college/Associate degree | Probably get a vaccine | 0.03651091 |
| Black | 55-64 | Male | Incomplete college/Associate degree | Probably NOT get a vaccine | 0.00437121 |
| Black | 55-64 | Male | Incomplete college/Associate degree | Definitely NOT get a vaccine | 0.00674703 |
| Black | 55-64 | Male | Incomplete college/Associate degree | Already vaccinated | 0.17262389 |
| Black | 55-64 | Male | Bachelor degree or higher | Definitely get a vaccine | 0.04867214 |
| Black | 55-64 | Male | Bachelor degree or higher | Probably get a vaccine | 0.01035086 |
| Black | 55-64 | Male | Bachelor degree or higher | Probably NOT get a vaccine | 0.00288946 |
| Black | 55-64 | Male | Bachelor degree or higher | Definitely NOT get a vaccine | 0.00398828 |
| Black | 55-64 | Male | Bachelor degree or higher | Already vaccinated | 0.13965130 |
| Black | 55-64 | Female | High school or less | Definitely get a vaccine | 0.12730308 |
| Black | 55-64 | Female | High school or less | Probably get a vaccine | 0.09843513 |
| Black | 55-64 | Female | High school or less | Probably NOT get a vaccine | 0.01891362 |
| Black | 55-64 | Female | High school or less | Definitely NOT get a vaccine | 0.01702204 |
| Black | 55-64 | Female | High school or less | Already vaccinated | 0.20194656 |

| | | | | | |
|---|---|---|---|---|---|
| Black | 55-64 | Female | Incomplete college/Associate degree | Definitely get a vaccine | 0.09857206 |
| Black | 55-64 | Female | Incomplete college/Associate degree | Probably get a vaccine | 0.06397370 |
| Black | 55-64 | Female | Incomplete college/Associate degree | Probably NOT get a vaccine | 0.02709497 |
| Black | 55-64 | Female | Incomplete college/Associate degree | Definitely NOT get a vaccine | 0.01041587 |
| Black | 55-64 | Female | Incomplete college/Associate degree | Already vaccinated | 0.21106210 |
| Black | 55-64 | Female | Bachelor degree or higher | Definitely get a vaccine | 0.05444974 |
| Black | 55-64 | Female | Bachelor degree or higher | Probably get a vaccine | 0.03155396 |
| Black | 55-64 | Female | Bachelor degree or higher | Probably NOT get a vaccine | 0.00737329 |
| Black | 55-64 | Female | Bachelor degree or higher | Definitely NOT get a vaccine | 0.00657467 |
| Black | 55-64 | Female | Bachelor degree or higher | Already vaccinated | 0.22033820 |
| Black | 65+ | Male | High school or less | Definitely get a vaccine | 0.02850046 |
| Black | 65+ | Male | High school or less | Probably get a vaccine | 0.02460403 |
| Black | 65+ | Male | High school or less | Probably NOT get a vaccine | 0.00210334 |
| Black | 65+ | Male | High school or less | Definitely NOT get a vaccine | 0.03370555 |
| Black | 65+ | Male | High school or less | Already vaccinated | 0.35494372 |
| Black | 65+ | Male | Incomplete college/Associate degree | Definitely get a vaccine | 0.02465111 |
| Black | 65+ | Male | Incomplete college/Associate degree | Probably get a vaccine | 0.01105626 |
| Black | 65+ | Male | Incomplete college/Associate degree | Probably NOT get a vaccine | 0.00153188 |
| Black | 65+ | Male | Incomplete college/Associate degree | Definitely NOT get a vaccine | 0.00303603 |
| Black | 65+ | Male | Incomplete college/Associate degree | Already vaccinated | 0.19647512 |
| Black | 65+ | Male | Bachelor degree or higher | Definitely get a vaccine | 0.01729118 |
| Black | 65+ | Male | Bachelor degree or higher | Probably get a vaccine | 0.00309439 |
| Black | 65+ | Male | Bachelor degree or higher | Probably NOT get a vaccine | 0.00049318 |
| Black | 65+ | Male | Bachelor degree or higher | Definitely NOT get a vaccine | 0.00067654 |
| Black | 65+ | Male | Bachelor degree or higher | Already vaccinated | 0.17602876 |
| Black | 65+ | Female | High school or less | Definitely get a vaccine | 0.12069507 |
| Black | 65+ | Female | High school or less | Probably get a vaccine | 0.01418060 |
| Black | 65+ | Female | High school or less | Probably NOT get a vaccine | 0.05566464 |
| Black | 65+ | Female | High school or less | Definitely NOT get a vaccine | 0.00510152 |
| Black | 65+ | Female | High school or less | Already vaccinated | 0.41330323 |
| Black | 65+ | Female | Incomplete college/Associate degree | Definitely get a vaccine | 0.03523853 |
| Black | 65+ | Female | Incomplete college/Associate degree | Probably get a vaccine | 0.01130142 |
| Black | 65+ | Female | Incomplete college/Associate degree | Probably NOT get a vaccine | 0.02107424 |
| Black | 65+ | Female | Incomplete college/Associate degree | Definitely NOT get a vaccine | 0.00313218 |
| Black | 65+ | Female | Incomplete college/Associate degree | Already vaccinated | 0.28064644 |
| Black | 65+ | Female | Bachelor degree or higher | Definitely get a vaccine | 0.01852849 |
| Black | 65+ | Female | Bachelor degree or higher | Probably get a vaccine | 0.00616889 |
| Black | 65+ | Female | Bachelor degree or higher | Probably NOT get a vaccine | 0.00309576 |
| Black | 65+ | Female | Bachelor degree or higher | Definitely NOT get a vaccine | 0.00262021 |
| Black | 65+ | Female | Bachelor degree or higher | Already vaccinated | 0.25469264 |
| Asian | 18-24 | Male | High school or less | Definitely get a vaccine | 0.08925343 |
| Asian | 18-24 | Male | High school or less | Probably get a vaccine | 0.05172164 |
| Asian | 18-24 | Male | High school or less | Probably NOT get a vaccine | 0.00514875 |
| Asian | 18-24 | Male | High school or less | Definitely NOT get a vaccine | 0.00957360 |
| Asian | 18-24 | Male | High school or less | Already vaccinated | 0.02536161 |
| Asian | 18-24 | Male | Incomplete college/Associate degree | Definitely get a vaccine | 0.05417718 |

| | | | | | |
|---|---|---|---|---|---|
| Asian | 18-24 | Male | Incomplete college/Associate degree | Probably get a vaccine | 0.01667725 |
| Asian | 18-24 | Male | Incomplete college/Associate degree | Probably NOT get a vaccine | 0.01107464 |
| Asian | 18-24 | Male | Incomplete college/Associate degree | Definitely NOT get a vaccine | 0.00000000 |
| Asian | 18-24 | Male | Incomplete college/Associate degree | Already vaccinated | 0.03983258 |
| Asian | 18-24 | Male | Bachelor degree or higher | Definitely get a vaccine | 0.03088524 |
| Asian | 18-24 | Male | Bachelor degree or higher | Probably get a vaccine | 0.00154429 |
| Asian | 18-24 | Male | Bachelor degree or higher | Probably NOT get a vaccine | 0.00077361 |
| Asian | 18-24 | Male | Bachelor degree or higher | Definitely NOT get a vaccine | 0.00000000 |
| Asian | 18-24 | Male | Bachelor degree or higher | Already vaccinated | 0.02231143 |
| Asian | 18-24 | Female | High school or less | Definitely get a vaccine | 0.03999595 |
| Asian | 18-24 | Female | High school or less | Probably get a vaccine | 0.01543754 |
| Asian | 18-24 | Female | High school or less | Probably NOT get a vaccine | 0.01152362 |
| Asian | 18-24 | Female | High school or less | Definitely NOT get a vaccine | 0.00000000 |
| Asian | 18-24 | Female | High school or less | Already vaccinated | 0.02519220 |
| Asian | 18-24 | Female | Incomplete college/Associate degree | Definitely get a vaccine | 0.07995502 |
| Asian | 18-24 | Female | Incomplete college/Associate degree | Probably get a vaccine | 0.01806746 |
| Asian | 18-24 | Female | Incomplete college/Associate degree | Probably NOT get a vaccine | 0.01214414 |
| Asian | 18-24 | Female | Incomplete college/Associate degree | Definitely NOT get a vaccine | 0.00000000 |
| Asian | 18-24 | Female | Incomplete college/Associate degree | Already vaccinated | 0.06719825 |
| Asian | 18-24 | Female | Bachelor degree or higher | Definitely get a vaccine | 0.01799504 |
| Asian | 18-24 | Female | Bachelor degree or higher | Probably get a vaccine | 0.01015665 |
| Asian | 18-24 | Female | Bachelor degree or higher | Probably NOT get a vaccine | 0.00155166 |
| Asian | 18-24 | Female | Bachelor degree or higher | Definitely NOT get a vaccine | 0.00000000 |
| Asian | 18-24 | Female | Bachelor degree or higher | Already vaccinated | 0.06306288 |
| Asian | 25-34 | Male | High school or less | Definitely get a vaccine | 0.02730611 |
| Asian | 25-34 | Male | High school or less | Probably get a vaccine | 0.02823729 |
| Asian | 25-34 | Male | High school or less | Probably NOT get a vaccine | 0.00000000 |
| Asian | 25-34 | Male | High school or less | Definitely NOT get a vaccine | 0.00387910 |
| Asian | 25-34 | Male | High school or less | Already vaccinated | 0.04056692 |
| Asian | 25-34 | Male | Incomplete college/Associate degree | Definitely get a vaccine | 0.03521256 |
| Asian | 25-34 | Male | Incomplete college/Associate degree | Probably get a vaccine | 0.04019436 |
| Asian | 25-34 | Male | Incomplete college/Associate degree | Probably NOT get a vaccine | 0.00265164 |
| Asian | 25-34 | Male | Incomplete college/Associate degree | Definitely NOT get a vaccine | 0.00197536 |
| Asian | 25-34 | Male | Incomplete college/Associate degree | Already vaccinated | 0.02133662 |
| Asian | 25-34 | Male | Bachelor degree or higher | Definitely get a vaccine | 0.14514023 |
| Asian | 25-34 | Male | Bachelor degree or higher | Probably get a vaccine | 0.03017174 |
| Asian | 25-34 | Male | Bachelor degree or higher | Probably NOT get a vaccine | 0.01254402 |
| Asian | 25-34 | Male | Bachelor degree or higher | Definitely NOT get a vaccine | 0.00033734 |
| Asian | 25-34 | Male | Bachelor degree or higher | Already vaccinated | 0.14795685 |
| Asian | 25-34 | Female | High school or less | Definitely get a vaccine | 0.06887820 |
| Asian | 25-34 | Female | High school or less | Probably get a vaccine | 0.01592830 |
| Asian | 25-34 | Female | High school or less | Probably NOT get a vaccine | 0.00752822 |
| Asian | 25-34 | Female | High school or less | Definitely NOT get a vaccine | 0.00401701 |
| Asian | 25-34 | Female | High school or less | Already vaccinated | 0.03726279 |
| Asian | 25-34 | Female | Incomplete college/Associate degree | Definitely get a vaccine | 0.02991411 |
| Asian | 25-34 | Female | Incomplete college/Associate degree | Probably get a vaccine | 0.01958547 |

| | | | | | |
|---|---|---|---|---|---|
| Asian | 25-34 | Female | Incomplete college/Associate degree | Probably NOT get a vaccine | 0.00092931 |
| Asian | 25-34 | Female | Incomplete college/Associate degree | Definitely NOT get a vaccine | 0.00000000 |
| Asian | 25-34 | Female | Incomplete college/Associate degree | Already vaccinated | 0.03250938 |
| Asian | 25-34 | Female | Bachelor degree or higher | Definitely get a vaccine | 0.15728934 |
| Asian | 25-34 | Female | Bachelor degree or higher | Probably get a vaccine | 0.02950131 |
| Asian | 25-34 | Female | Bachelor degree or higher | Probably NOT get a vaccine | 0.02915342 |
| Asian | 25-34 | Female | Bachelor degree or higher | Definitely NOT get a vaccine | 0.00514116 |
| Asian | 25-34 | Female | Bachelor degree or higher | Already vaccinated | 0.17112930 |
| Asian | 35-44 | Male | High school or less | Definitely get a vaccine | 0.06076729 |
| Asian | 35-44 | Male | High school or less | Probably get a vaccine | 0.05140866 |
| Asian | 35-44 | Male | High school or less | Probably NOT get a vaccine | 0.00277778 |
| Asian | 35-44 | Male | High school or less | Definitely NOT get a vaccine | 0.00000000 |
| Asian | 35-44 | Male | High school or less | Already vaccinated | 0.03048573 |
| Asian | 35-44 | Male | Incomplete college/Associate degree | Definitely get a vaccine | 0.01820107 |
| Asian | 35-44 | Male | Incomplete college/Associate degree | Probably get a vaccine | 0.01286178 |
| Asian | 35-44 | Male | Incomplete college/Associate degree | Probably NOT get a vaccine | 0.00547024 |
| Asian | 35-44 | Male | Incomplete college/Associate degree | Definitely NOT get a vaccine | 0.00133333 |
| Asian | 35-44 | Male | Incomplete college/Associate degree | Already vaccinated | 0.02919496 |
| Asian | 35-44 | Male | Bachelor degree or higher | Definitely get a vaccine | 0.14545323 |
| Asian | 35-44 | Male | Bachelor degree or higher | Probably get a vaccine | 0.02498438 |
| Asian | 35-44 | Male | Bachelor degree or higher | Probably NOT get a vaccine | 0.00712503 |
| Asian | 35-44 | Male | Bachelor degree or higher | Definitely NOT get a vaccine | 0.00180215 |
| Asian | 35-44 | Male | Bachelor degree or higher | Already vaccinated | 0.19059734 |
| Asian | 35-44 | Female | High school or less | Definitely get a vaccine | 0.04444953 |
| Asian | 35-44 | Female | High school or less | Probably get a vaccine | 0.03208350 |
| Asian | 35-44 | Female | High school or less | Probably NOT get a vaccine | 0.04197700 |
| Asian | 35-44 | Female | High school or less | Definitely NOT get a vaccine | 0.00213067 |
| Asian | 35-44 | Female | High school or less | Already vaccinated | 0.02019734 |
| Asian | 35-44 | Female | Incomplete college/Associate degree | Definitely get a vaccine | 0.02132005 |
| Asian | 35-44 | Female | Incomplete college/Associate degree | Probably get a vaccine | 0.01648900 |
| Asian | 35-44 | Female | Incomplete college/Associate degree | Probably NOT get a vaccine | 0.00595041 |
| Asian | 35-44 | Female | Incomplete college/Associate degree | Definitely NOT get a vaccine | 0.00315473 |
| Asian | 35-44 | Female | Incomplete college/Associate degree | Already vaccinated | 0.06090214 |
| Asian | 35-44 | Female | Bachelor degree or higher | Definitely get a vaccine | 0.10364696 |
| Asian | 35-44 | Female | Bachelor degree or higher | Probably get a vaccine | 0.03620420 |
| Asian | 35-44 | Female | Bachelor degree or higher | Probably NOT get a vaccine | 0.01210910 |
| Asian | 35-44 | Female | Bachelor degree or higher | Definitely NOT get a vaccine | 0.00608550 |
| Asian | 35-44 | Female | Bachelor degree or higher | Already vaccinated | 0.17777827 |
| Asian | 45-54 | Male | High school or less | Definitely get a vaccine | 0.10716453 |
| Asian | 45-54 | Male | High school or less | Probably get a vaccine | 0.00985087 |
| Asian | 45-54 | Male | High school or less | Probably NOT get a vaccine | 0.00000000 |
| Asian | 45-54 | Male | High school or less | Definitely NOT get a vaccine | 0.01794109 |
| Asian | 45-54 | Male | High school or less | Already vaccinated | 0.04811753 |
| Asian | 45-54 | Male | Incomplete college/Associate degree | Definitely get a vaccine | 0.05046298 |
| Asian | 45-54 | Male | Incomplete college/Associate degree | Probably get a vaccine | 0.00104424 |
| Asian | 45-54 | Male | Incomplete college/Associate degree | Probably NOT get a vaccine | 0.00221265 |

| | | | | | |
|---|---|---|---|---|---|
| Asian | 45-54 | Male | Incomplete college/Associate degree | Definitely NOT get a vaccine | 0.00000000 |
| Asian | 45-54 | Male | Incomplete college/Associate degree | Already vaccinated | 0.03858042 |
| Asian | 45-54 | Male | Bachelor degree or higher | Definitely get a vaccine | 0.08056107 |
| Asian | 45-54 | Male | Bachelor degree or higher | Probably get a vaccine | 0.02463886 |
| Asian | 45-54 | Male | Bachelor degree or higher | Probably NOT get a vaccine | 0.00107174 |
| Asian | 45-54 | Male | Bachelor degree or higher | Definitely NOT get a vaccine | 0.00255383 |
| Asian | 45-54 | Male | Bachelor degree or higher | Already vaccinated | 0.15283674 |
| Asian | 45-54 | Female | High school or less | Definitely get a vaccine | 0.06421435 |
| Asian | 45-54 | Female | High school or less | Probably get a vaccine | 0.00397683 |
| Asian | 45-54 | Female | High school or less | Probably NOT get a vaccine | 0.00082658 |
| Asian | 45-54 | Female | High school or less | Definitely NOT get a vaccine | 0.00513027 |
| Asian | 45-54 | Female | High school or less | Already vaccinated | 0.06932677 |
| Asian | 45-54 | Female | Incomplete college/Associate degree | Definitely get a vaccine | 0.05216686 |
| Asian | 45-54 | Female | Incomplete college/Associate degree | Probably get a vaccine | 0.00874338 |
| Asian | 45-54 | Female | Incomplete college/Associate degree | Probably NOT get a vaccine | 0.00089337 |
| Asian | 45-54 | Female | Incomplete college/Associate degree | Definitely NOT get a vaccine | 0.00158509 |
| Asian | 45-54 | Female | Incomplete college/Associate degree | Already vaccinated | 0.07866634 |
| Asian | 45-54 | Female | Bachelor degree or higher | Definitely get a vaccine | 0.06516548 |
| Asian | 45-54 | Female | Bachelor degree or higher | Probably get a vaccine | 0.01226359 |
| Asian | 45-54 | Female | Bachelor degree or higher | Probably NOT get a vaccine | 0.00606057 |
| Asian | 45-54 | Female | Bachelor degree or higher | Definitely NOT get a vaccine | 0.00276439 |
| Asian | 45-54 | Female | Bachelor degree or higher | Already vaccinated | 0.15696833 |
| Asian | 55-64 | Male | High school or less | Definitely get a vaccine | 0.06129467 |
| Asian | 55-64 | Male | High school or less | Probably get a vaccine | 0.02789364 |
| Asian | 55-64 | Male | High school or less | Probably NOT get a vaccine | 0.00000000 |
| Asian | 55-64 | Male | High school or less | Definitely NOT get a vaccine | 0.03074940 |
| Asian | 55-64 | Male | High school or less | Already vaccinated | 0.08722611 |
| Asian | 55-64 | Male | Incomplete college/Associate degree | Definitely get a vaccine | 0.03167597 |
| Asian | 55-64 | Male | Incomplete college/Associate degree | Probably get a vaccine | 0.00757007 |
| Asian | 55-64 | Male | Incomplete college/Associate degree | Probably NOT get a vaccine | 0.00231137 |
| Asian | 55-64 | Male | Incomplete college/Associate degree | Definitely NOT get a vaccine | 0.00083119 |
| Asian | 55-64 | Male | Incomplete college/Associate degree | Already vaccinated | 0.05368098 |
| Asian | 55-64 | Male | Bachelor degree or higher | Definitely get a vaccine | 0.06896146 |
| Asian | 55-64 | Male | Bachelor degree or higher | Probably get a vaccine | 0.00859401 |
| Asian | 55-64 | Male | Bachelor degree or higher | Probably NOT get a vaccine | 0.00280690 |
| Asian | 55-64 | Male | Bachelor degree or higher | Definitely NOT get a vaccine | 0.00360935 |
| Asian | 55-64 | Male | Bachelor degree or higher | Already vaccinated | 0.12885049 |
| Asian | 55-64 | Female | High school or less | Definitely get a vaccine | 0.03594074 |
| Asian | 55-64 | Female | High school or less | Probably get a vaccine | 0.00590746 |
| Asian | 55-64 | Female | High school or less | Probably NOT get a vaccine | 0.00122615 |
| Asian | 55-64 | Female | High school or less | Definitely NOT get a vaccine | 0.00283710 |
| Asian | 55-64 | Female | High school or less | Already vaccinated | 0.04762214 |
| Asian | 55-64 | Female | Incomplete college/Associate degree | Definitely get a vaccine | 0.04800200 |
| Asian | 55-64 | Female | Incomplete college/Associate degree | Probably get a vaccine | 0.00607689 |
| Asian | 55-64 | Female | Incomplete college/Associate degree | Probably NOT get a vaccine | 0.00182600 |
| Asian | 55-64 | Female | Incomplete college/Associate degree | Definitely NOT get a vaccine | 0.00077741 |

| | | | | | |
|---|---|---|---|---|---|
| Asian | 55-64 | Female | Incomplete college/Associate degree | Already vaccinated | 0.05289338 |
| Asian | 55-64 | Female | Bachelor degree or higher | Definitely get a vaccine | 0.07205372 |
| Asian | 55-64 | Female | Bachelor degree or higher | Probably get a vaccine | 0.00633430 |
| Asian | 55-64 | Female | Bachelor degree or higher | Probably NOT get a vaccine | 0.00148692 |
| Asian | 55-64 | Female | Bachelor degree or higher | Definitely NOT get a vaccine | 0.00420438 |
| Asian | 55-64 | Female | Bachelor degree or higher | Already vaccinated | 0.17571425 |
| Asian | 65+ | Male | High school or less | Definitely get a vaccine | 0.02843347 |
| Asian | 65+ | Male | High school or less | Probably get a vaccine | 0.00792447 |
| Asian | 65+ | Male | High school or less | Probably NOT get a vaccine | 0.00121173 |
| Asian | 65+ | Male | High school or less | Definitely NOT get a vaccine | 0.00562747 |
| Asian | 65+ | Male | High school or less | Already vaccinated | 0.11082763 |
| Asian | 65+ | Male | Incomplete college/Associate degree | Definitely get a vaccine | 0.00265349 |
| Asian | 65+ | Male | Incomplete college/Associate degree | Probably get a vaccine | 0.00005630 |
| Asian | 65+ | Male | Incomplete college/Associate degree | Probably NOT get a vaccine | 0.00000000 |
| Asian | 65+ | Male | Incomplete college/Associate degree | Definitely NOT get a vaccine | 0.00000000 |
| Asian | 65+ | Male | Incomplete college/Associate degree | Already vaccinated | 0.06814505 |
| Asian | 65+ | Male | Bachelor degree or higher | Definitely get a vaccine | 0.01674307 |
| Asian | 65+ | Male | Bachelor degree or higher | Probably get a vaccine | 0.00196322 |
| Asian | 65+ | Male | Bachelor degree or higher | Probably NOT get a vaccine | 0.00070353 |
| Asian | 65+ | Male | Bachelor degree or higher | Definitely NOT get a vaccine | 0.00729074 |
| Asian | 65+ | Male | Bachelor degree or higher | Already vaccinated | 0.17292570 |
| Asian | 65+ | Female | High school or less | Definitely get a vaccine | 0.04878502 |
| Asian | 65+ | Female | High school or less | Probably get a vaccine | 0.00072172 |
| Asian | 65+ | Female | High school or less | Probably NOT get a vaccine | 0.00048115 |
| Asian | 65+ | Female | High school or less | Definitely NOT get a vaccine | 0.00176244 |
| Asian | 65+ | Female | High school or less | Already vaccinated | 0.15986237 |
| Asian | 65+ | Female | Incomplete college/Associate degree | Definitely get a vaccine | 0.01266344 |
| Asian | 65+ | Female | Incomplete college/Associate degree | Probably get a vaccine | 0.00101169 |
| Asian | 65+ | Female | Incomplete college/Associate degree | Probably NOT get a vaccine | 0.00075256 |
| Asian | 65+ | Female | Incomplete college/Associate degree | Definitely NOT get a vaccine | 0.00115714 |
| Asian | 65+ | Female | Incomplete college/Associate degree | Already vaccinated | 0.07141849 |
| Asian | 65+ | Female | Bachelor degree or higher | Definitely get a vaccine | 0.00530137 |
| Asian | 65+ | Female | Bachelor degree or higher | Probably get a vaccine | 0.00299273 |
| Asian | 65+ | Female | Bachelor degree or higher | Probably NOT get a vaccine | 0.00551290 |
| Asian | 65+ | Female | Bachelor degree or higher | Definitely NOT get a vaccine | 0.00162664 |
| Asian | 65+ | Female | Bachelor degree or higher | Already vaccinated | 0.14017583 |
| Other | 18-24 | Male | High school or less | Definitely get a vaccine | 0.02134556 |
| Other | 18-24 | Male | High school or less | Probably get a vaccine | 0.03484817 |
| Other | 18-24 | Male | High school or less | Probably NOT get a vaccine | 0.03695260 |
| Other | 18-24 | Male | High school or less | Definitely NOT get a vaccine | 0.01833893 |
| Other | 18-24 | Male | High school or less | Already vaccinated | 0.01652596 |
| Other | 18-24 | Male | Incomplete college/Associate degree | Definitely get a vaccine | 0.13181543 |
| Other | 18-24 | Male | Incomplete college/Associate degree | Probably get a vaccine | 0.00927891 |
| Other | 18-24 | Male | Incomplete college/Associate degree | Probably NOT get a vaccine | 0.00472744 |
| Other | 18-24 | Male | Incomplete college/Associate degree | Definitely NOT get a vaccine | 0.01585518 |
| Other | 18-24 | Male | Incomplete college/Associate degree | Already vaccinated | 0.00712855 |

| | | | | | |
|---|---|---|---|---|---|
| Other | 18-24 | Male | Bachelor degree or higher | Definitely get a vaccine | 0.02276208 |
| Other | 18-24 | Male | Bachelor degree or higher | Probably get a vaccine | 0.00385456 |
| Other | 18-24 | Male | Bachelor degree or higher | Probably NOT get a vaccine | 0.00000000 |
| Other | 18-24 | Male | Bachelor degree or higher | Definitely NOT get a vaccine | 0.00000000 |
| Other | 18-24 | Male | Bachelor degree or higher | Already vaccinated | 0.00500574 |
| Other | 18-24 | Female | High school or less | Definitely get a vaccine | 0.09231015 |
| Other | 18-24 | Female | High school or less | Probably get a vaccine | 0.03309172 |
| Other | 18-24 | Female | High school or less | Probably NOT get a vaccine | 0.01209313 |
| Other | 18-24 | Female | High school or less | Definitely NOT get a vaccine | 0.02633066 |
| Other | 18-24 | Female | High school or less | Already vaccinated | 0.00833759 |
| Other | 18-24 | Female | Incomplete college/Associate degree | Definitely get a vaccine | 0.07724159 |
| Other | 18-24 | Female | Incomplete college/Associate degree | Probably get a vaccine | 0.02892457 |
| Other | 18-24 | Female | Incomplete college/Associate degree | Probably NOT get a vaccine | 0.02408120 |
| Other | 18-24 | Female | Incomplete college/Associate degree | Definitely NOT get a vaccine | 0.00716477 |
| Other | 18-24 | Female | Incomplete college/Associate degree | Already vaccinated | 0.04024077 |
| Other | 18-24 | Female | Bachelor degree or higher | Definitely get a vaccine | 0.00539392 |
| Other | 18-24 | Female | Bachelor degree or higher | Probably get a vaccine | 0.00008376 |
| Other | 18-24 | Female | Bachelor degree or higher | Probably NOT get a vaccine | 0.00000000 |
| Other | 18-24 | Female | Bachelor degree or higher | Definitely NOT get a vaccine | 0.00088037 |
| Other | 18-24 | Female | Bachelor degree or higher | Already vaccinated | 0.01093880 |
| Other | 25-34 | Male | High school or less | Definitely get a vaccine | 0.12776446 |
| Other | 25-34 | Male | High school or less | Probably get a vaccine | 0.07900280 |
| Other | 25-34 | Male | High school or less | Probably NOT get a vaccine | 0.06186034 |
| Other | 25-34 | Male | High school or less | Definitely NOT get a vaccine | 0.01137841 |
| Other | 25-34 | Male | High school or less | Already vaccinated | 0.03519610 |
| Other | 25-34 | Male | Incomplete college/Associate degree | Definitely get a vaccine | 0.05805614 |
| Other | 25-34 | Male | Incomplete college/Associate degree | Probably get a vaccine | 0.01644711 |
| Other | 25-34 | Male | Incomplete college/Associate degree | Probably NOT get a vaccine | 0.01484882 |
| Other | 25-34 | Male | Incomplete college/Associate degree | Definitely NOT get a vaccine | 0.01232191 |
| Other | 25-34 | Male | Incomplete college/Associate degree | Already vaccinated | 0.02560084 |
| Other | 25-34 | Male | Bachelor degree or higher | Definitely get a vaccine | 0.05096770 |
| Other | 25-34 | Male | Bachelor degree or higher | Probably get a vaccine | 0.02045735 |
| Other | 25-34 | Male | Bachelor degree or higher | Probably NOT get a vaccine | 0.00743444 |
| Other | 25-34 | Male | Bachelor degree or higher | Definitely NOT get a vaccine | 0.01098614 |
| Other | 25-34 | Male | Bachelor degree or higher | Already vaccinated | 0.05757666 |
| Other | 25-34 | Female | High school or less | Definitely get a vaccine | 0.04382529 |
| Other | 25-34 | Female | High school or less | Probably get a vaccine | 0.06850471 |
| Other | 25-34 | Female | High school or less | Probably NOT get a vaccine | 0.03456258 |
| Other | 25-34 | Female | High school or less | Definitely NOT get a vaccine | 0.02769851 |
| Other | 25-34 | Female | High school or less | Already vaccinated | 0.01326296 |
| Other | 25-34 | Female | Incomplete college/Associate degree | Definitely get a vaccine | 0.07752993 |
| Other | 25-34 | Female | Incomplete college/Associate degree | Probably get a vaccine | 0.03673761 |
| Other | 25-34 | Female | Incomplete college/Associate degree | Probably NOT get a vaccine | 0.03803600 |
| Other | 25-34 | Female | Incomplete college/Associate degree | Definitely NOT get a vaccine | 0.02936455 |
| Other | 25-34 | Female | Incomplete college/Associate degree | Already vaccinated | 0.05876022 |
| Other | 25-34 | Female | Bachelor degree or higher | Definitely get a vaccine | 0.04771763 |

| | | | | | |
|---|---|---|---|---|---|
| Other | 25-34 | Female | Bachelor degree or higher | Probably get a vaccine | 0.01873126 |
| Other | 25-34 | Female | Bachelor degree or higher | Probably NOT get a vaccine | 0.03577077 |
| Other | 25-34 | Female | Bachelor degree or higher | Definitely NOT get a vaccine | 0.00675523 |
| Other | 25-34 | Female | Bachelor degree or higher | Already vaccinated | 0.10785677 |
| Other | 35-44 | Male | High school or less | Definitely get a vaccine | 0.08474768 |
| Other | 35-44 | Male | High school or less | Probably get a vaccine | 0.04134865 |
| Other | 35-44 | Male | High school or less | Probably NOT get a vaccine | 0.05452959 |
| Other | 35-44 | Male | High school or less | Definitely NOT get a vaccine | 0.04386125 |
| Other | 35-44 | Male | High school or less | Already vaccinated | 0.05940614 |
| Other | 35-44 | Male | Incomplete college/Associate degree | Definitely get a vaccine | 0.04576201 |
| Other | 35-44 | Male | Incomplete college/Associate degree | Probably get a vaccine | 0.02808035 |
| Other | 35-44 | Male | Incomplete college/Associate degree | Probably NOT get a vaccine | 0.00842230 |
| Other | 35-44 | Male | Incomplete college/Associate degree | Definitely NOT get a vaccine | 0.02271898 |
| Other | 35-44 | Male | Incomplete college/Associate degree | Already vaccinated | 0.07699941 |
| Other | 35-44 | Male | Bachelor degree or higher | Definitely get a vaccine | 0.02270526 |
| Other | 35-44 | Male | Bachelor degree or higher | Probably get a vaccine | 0.00743178 |
| Other | 35-44 | Male | Bachelor degree or higher | Probably NOT get a vaccine | 0.02267151 |
| Other | 35-44 | Male | Bachelor degree or higher | Definitely NOT get a vaccine | 0.00992088 |
| Other | 35-44 | Male | Bachelor degree or higher | Already vaccinated | 0.06387236 |
| Other | 35-44 | Female | High school or less | Definitely get a vaccine | 0.04523140 |
| Other | 35-44 | Female | High school or less | Probably get a vaccine | 0.07287856 |
| Other | 35-44 | Female | High school or less | Probably NOT get a vaccine | 0.02514067 |
| Other | 35-44 | Female | High school or less | Definitely NOT get a vaccine | 0.05906947 |
| Other | 35-44 | Female | High school or less | Already vaccinated | 0.06503431 |
| Other | 35-44 | Female | Incomplete college/Associate degree | Definitely get a vaccine | 0.04828341 |
| Other | 35-44 | Female | Incomplete college/Associate degree | Probably get a vaccine | 0.02150047 |
| Other | 35-44 | Female | Incomplete college/Associate degree | Probably NOT get a vaccine | 0.03593946 |
| Other | 35-44 | Female | Incomplete college/Associate degree | Definitely NOT get a vaccine | 0.03758369 |
| Other | 35-44 | Female | Incomplete college/Associate degree | Already vaccinated | 0.07525337 |
| Other | 35-44 | Female | Bachelor degree or higher | Definitely get a vaccine | 0.03592234 |
| Other | 35-44 | Female | Bachelor degree or higher | Probably get a vaccine | 0.01493844 |
| Other | 35-44 | Female | Bachelor degree or higher | Probably NOT get a vaccine | 0.01784177 |
| Other | 35-44 | Female | Bachelor degree or higher | Definitely NOT get a vaccine | 0.01219948 |
| Other | 35-44 | Female | Bachelor degree or higher | Already vaccinated | 0.07802971 |
| Other | 45-54 | Male | High school or less | Definitely get a vaccine | 0.08802198 |
| Other | 45-54 | Male | High school or less | Probably get a vaccine | 0.03502189 |
| Other | 45-54 | Male | High school or less | Probably NOT get a vaccine | 0.07402769 |
| Other | 45-54 | Male | High school or less | Definitely NOT get a vaccine | 0.05621023 |
| Other | 45-54 | Male | High school or less | Already vaccinated | 0.13367274 |
| Other | 45-54 | Male | Incomplete college/Associate degree | Definitely get a vaccine | 0.00839404 |
| Other | 45-54 | Male | Incomplete college/Associate degree | Probably get a vaccine | 0.02700370 |
| Other | 45-54 | Male | Incomplete college/Associate degree | Probably NOT get a vaccine | 0.00527595 |
| Other | 45-54 | Male | Incomplete college/Associate degree | Definitely NOT get a vaccine | 0.01321674 |
| Other | 45-54 | Male | Incomplete college/Associate degree | Already vaccinated | 0.05811513 |
| Other | 45-54 | Male | Bachelor degree or higher | Definitely get a vaccine | 0.02790177 |
| Other | 45-54 | Male | Bachelor degree or higher | Probably get a vaccine | 0.00501501 |

| | | | | | |
|---|---|---|---|---|---|
| Other | 45-54 | Male | Bachelor degree or higher | Probably NOT get a vaccine | 0.01020867 |
| Other | 45-54 | Male | Bachelor degree or higher | Definitely NOT get a vaccine | 0.01779168 |
| Other | 45-54 | Male | Bachelor degree or higher | Already vaccinated | 0.04079571 |
| Other | 45-54 | Female | High school or less | Definitely get a vaccine | 0.07911984 |
| Other | 45-54 | Female | High school or less | Probably get a vaccine | 0.03025230 |
| Other | 45-54 | Female | High school or less | Probably NOT get a vaccine | 0.02096736 |
| Other | 45-54 | Female | High school or less | Definitely NOT get a vaccine | 0.01936923 |
| Other | 45-54 | Female | High school or less | Already vaccinated | 0.08342101 |
| Other | 45-54 | Female | Incomplete college/Associate degree | Definitely get a vaccine | 0.02646384 |
| Other | 45-54 | Female | Incomplete college/Associate degree | Probably get a vaccine | 0.01233945 |
| Other | 45-54 | Female | Incomplete college/Associate degree | Probably NOT get a vaccine | 0.01221315 |
| Other | 45-54 | Female | Incomplete college/Associate degree | Definitely NOT get a vaccine | 0.02329923 |
| Other | 45-54 | Female | Incomplete college/Associate degree | Already vaccinated | 0.05832196 |
| Other | 45-54 | Female | Bachelor degree or higher | Definitely get a vaccine | 0.01531184 |
| Other | 45-54 | Female | Bachelor degree or higher | Probably get a vaccine | 0.00782408 |
| Other | 45-54 | Female | Bachelor degree or higher | Probably NOT get a vaccine | 0.00359570 |
| Other | 45-54 | Female | Bachelor degree or higher | Definitely NOT get a vaccine | 0.00559669 |
| Other | 45-54 | Female | Bachelor degree or higher | Already vaccinated | 0.06362495 |
| Other | 55-64 | Male | High school or less | Definitely get a vaccine | 0.07095152 |
| Other | 55-64 | Male | High school or less | Probably get a vaccine | 0.01693015 |
| Other | 55-64 | Male | High school or less | Probably NOT get a vaccine | 0.04932473 |
| Other | 55-64 | Male | High school or less | Definitely NOT get a vaccine | 0.01437207 |
| Other | 55-64 | Male | High school or less | Already vaccinated | 0.06810377 |
| Other | 55-64 | Male | Incomplete college/Associate degree | Definitely get a vaccine | 0.02758296 |
| Other | 55-64 | Male | Incomplete college/Associate degree | Probably get a vaccine | 0.01715118 |
| Other | 55-64 | Male | Incomplete college/Associate degree | Probably NOT get a vaccine | 0.02807585 |
| Other | 55-64 | Male | Incomplete college/Associate degree | Definitely NOT get a vaccine | 0.00918048 |
| Other | 55-64 | Male | Incomplete college/Associate degree | Already vaccinated | 0.04021316 |
| Other | 55-64 | Male | Bachelor degree or higher | Definitely get a vaccine | 0.01018427 |
| Other | 55-64 | Male | Bachelor degree or higher | Probably get a vaccine | 0.00348903 |
| Other | 55-64 | Male | Bachelor degree or higher | Probably NOT get a vaccine | 0.01075005 |
| Other | 55-64 | Male | Bachelor degree or higher | Definitely NOT get a vaccine | 0.00549751 |
| Other | 55-64 | Male | Bachelor degree or higher | Already vaccinated | 0.02429029 |
| Other | 55-64 | Female | High school or less | Definitely get a vaccine | 0.03847726 |
| Other | 55-64 | Female | High school or less | Probably get a vaccine | 0.03885213 |
| Other | 55-64 | Female | High school or less | Probably NOT get a vaccine | 0.00281373 |
| Other | 55-64 | Female | High school or less | Definitely NOT get a vaccine | 0.01573796 |
| Other | 55-64 | Female | High school or less | Already vaccinated | 0.07249836 |
| Other | 55-64 | Female | Incomplete college/Associate degree | Definitely get a vaccine | 0.04025226 |
| Other | 55-64 | Female | Incomplete college/Associate degree | Probably get a vaccine | 0.02057520 |
| Other | 55-64 | Female | Incomplete college/Associate degree | Probably NOT get a vaccine | 0.01498319 |
| Other | 55-64 | Female | Incomplete college/Associate degree | Definitely NOT get a vaccine | 0.01011756 |
| Other | 55-64 | Female | Incomplete college/Associate degree | Already vaccinated | 0.04652537 |
| Other | 55-64 | Female | Bachelor degree or higher | Definitely get a vaccine | 0.01765683 |
| Other | 55-64 | Female | Bachelor degree or higher | Probably get a vaccine | 0.00622235 |
| Other | 55-64 | Female | Bachelor degree or higher | Probably NOT get a vaccine | 0.00186721 |

| | | | | | |
|---|---|---|---|---|---|
| Other | 55-64 | Female | Bachelor degree or higher | Definitely NOT get a vaccine | 0.01312964 |
| Other | 55-64 | Female | Bachelor degree or higher | Already vaccinated | 0.06076447 |
| Other | 65+ | Male | High school or less | Definitely get a vaccine | 0.03506804 |
| Other | 65+ | Male | High school or less | Probably get a vaccine | 0.00710667 |
| Other | 65+ | Male | High school or less | Probably NOT get a vaccine | 0.00337353 |
| Other | 65+ | Male | High school or less | Definitely NOT get a vaccine | 0.04983411 |
| Other | 65+ | Male | High school or less | Already vaccinated | 0.11919767 |
| Other | 65+ | Male | Incomplete college/Associate degree | Definitely get a vaccine | 0.00624257 |
| Other | 65+ | Male | Incomplete college/Associate degree | Probably get a vaccine | 0.00445378 |
| Other | 65+ | Male | Incomplete college/Associate degree | Probably NOT get a vaccine | 0.00341884 |
| Other | 65+ | Male | Incomplete college/Associate degree | Definitely NOT get a vaccine | 0.00261004 |
| Other | 65+ | Male | Incomplete college/Associate degree | Already vaccinated | 0.05964538 |
| Other | 65+ | Male | Bachelor degree or higher | Definitely get a vaccine | 0.00243740 |
| Other | 65+ | Male | Bachelor degree or higher | Probably get a vaccine | 0.00479646 |
| Other | 65+ | Male | Bachelor degree or higher | Probably NOT get a vaccine | 0.00089168 |
| Other | 65+ | Male | Bachelor degree or higher | Definitely NOT get a vaccine | 0.01960215 |
| Other | 65+ | Male | Bachelor degree or higher | Already vaccinated | 0.06006961 |
| Other | 65+ | Female | High school or less | Definitely get a vaccine | 0.03970670 |
| Other | 65+ | Female | High school or less | Probably get a vaccine | 0.01987238 |
| Other | 65+ | Female | High school or less | Probably NOT get a vaccine | 0.00858451 |
| Other | 65+ | Female | High school or less | Definitely NOT get a vaccine | 0.01021777 |
| Other | 65+ | Female | High school or less | Already vaccinated | 0.12835556 |
| Other | 65+ | Female | Incomplete college/Associate degree | Definitely get a vaccine | 0.02009652 |
| Other | 65+ | Female | Incomplete college/Associate degree | Probably get a vaccine | 0.00649460 |
| Other | 65+ | Female | Incomplete college/Associate degree | Probably NOT get a vaccine | 0.00437677 |
| Other | 65+ | Female | Incomplete college/Associate degree | Definitely NOT get a vaccine | 0.00429728 |
| Other | 65+ | Female | Incomplete college/Associate degree | Already vaccinated | 0.07079219 |
| Other | 65+ | Female | Bachelor degree or higher | Definitely get a vaccine | 0.00763137 |
| Other | 65+ | Female | Bachelor degree or higher | Probably get a vaccine | 0.00315138 |
| Other | 65+ | Female | Bachelor degree or higher | Probably NOT get a vaccine | 0.00484022 |
| Other | 65+ | Female | Bachelor degree or higher | Definitely NOT get a vaccine | 0.00645835 |
| Other | 65+ | Female | Bachelor degree or higher | Already vaccinated | 0.06108241 |
